# Documented Dental Disease in Patients With *Staphylococcus aureus* Bacteremia

**DOI:** 10.64898/2026.10.18.26365833

**Authors:** Paddy Ssentongo, W. Frank Peacock, Jared C. Durieux, Christos Colovos

## Abstract

**Importance:** Clinicians often look for a dental source in *Staphylococcus aureus* bacteremia (SAB), but how often dental disease is found, compared with other bacteremias, is unclear.

**Objective:** To compare documented dental disease in adults with SAB, *Escherichia coli* bacteremia, and a general population cohort.

**Design, Setting, and Participants:** Retrospective cohort study in the Protégé database of 48 975 adults with bacteremia from January 1, 2015, through December 31, 2023, assigned by earliest in-window organism, and 81 945 general population adults.

**Exposures:** SAB, with *E coli* bacteremia as the primary comparator and the general population as the secondary comparator.

**Main Outcomes and Measures:** Documented dental disease from 7 days before through 30 days after index, defined by a qualifying dental procedure or disease-specific note text and adjusted for age, sex, race, and documentation intensity.

**Results:** Dental disease occurred in 528 of 16 435 patients with SAB (3.21%) and 179 of 24 942 patients with *E coli* bacteremia (0.72%). The adjusted odds ratio was 3.86 (95% CI, 3.23 to 4.63). By the pathology-only definition, disease occurred in 517 patients with SAB (3.15%) and 399 general population patients (0.49%). The adjusted odds ratio was 3.71 (95% CI, 3.08 to 4.46). Within SAB, rates were highest with prior infective endocarditis (9.8%), opioid use disorder (6.2%), injection drug use (5.3%), and prosthetic valve (5.0%). Prevalence ranged from 0.49% in the general population to 7.98% in other streptococcal bacteremia. Of 437 patients with SAB and an assignable first finding date, 333 (76.2%) had the finding after the index date.

**Conclusions and Relevance:** Dental disease was documented more often in SAB than in *E coli* bacteremia after adjustment. Most findings were first recorded after bacteremia was recognized, during SAB evaluation. These data do not show that dental disease caused the bacteremia.

**Key Points:** *Question:* Is dental disease documented more often in *Staphylococcus aureus* bacteremia (SAB) than in *Escherichia coli* bacteremia or a general-population cohort?

*Findings:* In 16 435 adults with SAB and 24 942 with *E coli* bacteremia, documented dental disease occurred in 3.21% and 0.72%, respectively. The adjusted odds ratio was 3.86 (95% CI, 3.23 to 4.63). Among SAB patients with an assignable date, 76.2% had the first finding after the index date.

*Meaning:* Dental disease was documented more often with SAB than with a nonoral bacteremia. This supports focused oral assessment during source evaluation, but does not establish the mouth as the source.

## Introduction

*Staphylococcus aureus* bacteremia (SAB) is common and often severe.^1^ It affected more than 119 000 patients in the United States in 2017.^2^ SAB can cause infective endocarditis, metastatic infection, relapse, and death.^2–4^ Management requires a careful search for the portal of entry and sites of spread.^5^ Endocarditis guidelines include the oral cavity among the portals that merit attention.^6,7^

The mouth is a recognized portal for viridans group streptococcal infection, in which dental procedures and daily oral activity can cause transient bacteremia.^8,9^ Its role in SAB is less clear. A systematic search for portals of entry in infective endocarditis found an oral or dental portal in only a minority of patients and included no contemporaneous nonoral bacteremia comparator.^10^ A dental finding in SAB may therefore reflect preexisting disease or the intensive search that follows SAB.^5^

Our objective was to determine whether dental disease was documented more often in SAB than in other bacteremias, which findings explained any excess, and whether disease was recorded before or after the source search began. We hypothesized that dental disease would be more common in SAB, mostly after the source search prompted closer oral assessment.

## Methods

### Data Source

This retrospective study used the Protégé research database (June 2026 release), a de-identified, longitudinal database that links electronic health records (EHR) with open medical and pharmacy claims. Protégé draws on routine-care EHR data from a network of 39 medium-to-large US health systems. Subsets of patients are also linked to radiology data and to dated records of death. The release contains 4 950 000 patients residing in all 50 US states and the District of Columbia, with 8.6 billion clinical events and 775 million clinical notes. For this study, we used data from January 1, 2015, to December 31, 2023 for the primary analyses. Database construction, scope, and de-identification are described in **eMethods 1** in the Supplement. Before delivery data was de-identified through expert determination. An external review by Bluebonnet Ethical Review determined that this activity is appropriately classified as Not Human Subjects Research (NHSR) and that IRB review is not required. Reporting follows RECORD and STROBE.^11,12^

### Study Population and Cohort Assignment

Cohorts were defined by prespecified organism-specific ICD-10-CM codes with a valid date, as in other large database studies.^13^ SAB used A41.01 and A41.02. *E coli* used A41.51. Other streptococcal sepsis used A40.8. Enterococcal sepsis used A41.81. Code A40.8 is a group code, not a species code, and likely includes mostly viridans group streptococci. Index was the earliest qualifying date in the window. Same day ties were assigned by the lowest code. Adults at index were included.

The primary window was January 1, 2015, through December 31, 2023, chosen because capture fell in later years. Vendor-shifted dates preserve within-patient intervals, so calendar labels are approximate. The full 2000 to 2025 span was used only in sensitivity analyses. The general population comparator was a 2 % deterministic hash sample. It required no qualifying bacteremia code at any time and a recorded birth year. Its first qualifying encounter was the pseudo index.

### Exposure and Outcome Definitions

The outcome was documented dental disease from 7 days before through 30 days after index. The locked definition, fixed before analysis, required a qualifying dental procedure or disease-specific text in a signed or addendum note. Qualifying text covered periapical lucency, radiolucency, infection, or abscess, dental, tooth, or odontogenic abscess, advanced or severe caries or carious lesions, retained roots, root tips, or fragments, and periodontal disease or periodontitis. Matching used word boundaries. A pathology-only variant excluded procedures and was used for population and cross-cohort comparisons, because procedures in the general population often reflect third-molar care in young adults. Nondental code families found in a pre-freeze audit were removed, and broad mentions or note type alone were not used. Code lists are in **eTable 1** in the Supplement.

### Covariates and Missing Data

Twenty comorbidities were defined from diagnosis codes in the 365 days before index. Age, sex, and race were measured at index (**eTable 1** in the Supplement). Documentation intensity was the number of signed or addendum notes in the window (0 to 1, 2 to 9, or 10 or more). Enterococcal characteristics are in **eTable 2** in the Supplement. Values were not imputed. Missing sex was analyzed as its own category (27 patients with SAB and 36 with *E coli* bacteremia). Race was White, Black, or other or unknown. Table 1 displays Asian separately; the model combined Asian with other or unknown.

**Table 1.** Baseline Characteristics by Cohort, 2015-2023.

| Characteristic | SAB | *E coli* BSI |  | Other strep. BSI |  | Enterococcal BSI |  | General population |  |
| --- | --- | --- | --- | --- | --- | --- | --- | --- | --- |
|  | (n=16 435) | (n=24 942) | SMD | (n=3097) | SMD | (n=4501) | SMD | (n=81 945) | SMD |
| **Demographics** |  |  |  |  |  |  |  |  |  |
| Age, mean (SD), y | 57.9 (17.2) | 65.2 (16.7) | 0.43 | 61.0 (16.7) | 0.18 | 66.8 (14.9) | 0.55 | 49.4 (18.2) | 0.48 |
| Male | 10 213 (62.1) | 9558 (38.3) | 0.49 | 1905 (61.5) | 0.01 | 2785 (61.9) | 0.01 | 34 261 (41.8) | 0.42 |
| White | 12 095 (73.6) | 17 113 (68.6) | 0.11 | 2166 (69.9) | 0.08 | 3206 (71.2) | 0.05 | 55 924 (68.2) | 0.12 |
| Black | 2281 (13.9) | 3520 (14.1) | 0.01 | 472 (15.2) | 0.04 | 647 (14.4) | 0.01 | 10 790 (13.2) | 0.02 |
| Asian | 510 (3.1) | 1319 (5.3) | 0.11 | 139 (4.5) | 0.07 | 229 (5.1) | 0.10 | 4098 (5.0) | 0.10 |
| Other or unknown | 1549 (9.4) | 2990 (12.0) | 0.08 | 320 (10.3) | 0.03 | 419 (9.3) | 0.00 | 11 133 (13.6) | 0.13 |
| **Cardiovascular history** |  |  |  |  |  |  |  |  |  |
| Heart failure | 2195 (13.4) | 2805 (11.2) | 0.06 | 427 (13.8) | 0.01 | 914 (20.3) | 0.19 | — | — |
| CAD or prior MI | 2425 (14.8) | 3657 (14.7) | 0.00 | 467 (15.1) | 0.01 | 1069 (23.8) | 0.23 | — | — |
| Cerebrovascular disease | 1171 (7.1) | 2053 (8.2) | 0.04 | 239 (7.7) | 0.02 | 566 (12.6) | 0.18 | — | — |
| Peripheral vascular disease | 1599 (9.7) | 2270 (9.1) | 0.02 | 331 (10.7) | 0.03 | 644 (14.3) | 0.14 | — | — |
| Native valve disease | 1111 (6.8) | 1365 (5.5) | 0.05 | 306 (9.9) | 0.11 | 571 (12.7) | 0.20 | — | — |
| Prosthetic valve | 319 (1.9) | 303 (1.2) | 0.06 | 130 (4.2) | 0.13 | 232 (5.2) | 0.17 | 103 (0.1) | 0.18 |
| Cardiac device | 589 (3.6) | 722 (2.9) | 0.04 | 99 (3.2) | 0.02 | 292 (6.5) | 0.13 | 208 (0.3) | 0.24 |
| Prior infective endocarditis | 379 (2.3) | 43 (0.2) | 0.19 | 78 (2.5) | 0.01 | 89 (2.0) | 0.02 | 11 (<0.1) | 0.22 |
| **Other comorbidities** |  |  |  |  |  |  |  |  |  |
| Diabetes | 4177 (25.4) | 5626 (22.6) | 0.07 | 780 (25.2) | 0.01 | 1287 (28.6) | 0.07 | — | — |
| Chronic kidney disease | 1937 (11.8) | 3402 (13.6) | 0.06 | 378 (12.2) | 0.01 | 876 (19.5) | 0.21 | — | — |
| End-stage kidney disease | 750 (4.6) | 446 (1.8) | 0.16 | 90 (2.9) | 0.09 | 227 (5.0) | 0.02 | 71 (0.1) | 0.30 |
| Liver disease | 978 (6.0) | 1777 (7.1) | 0.05 | 321 (10.4) | 0.16 | 530 (11.8) | 0.21 | — | — |
| COPD | 1615 (9.8) | 2169 (8.7) | 0.04 | 288 (9.3) | 0.02 | 575 (12.8) | 0.09 | — | — |
| Malignancy | 2011 (12.2) | 4348 (17.4) | 0.15 | 653 (21.1) | 0.24 | 1114 (24.8) | 0.33 | — | — |
| HIV or immunocompromise | 633 (3.9) | 1305 (5.2) | 0.07 | 190 (6.1) | 0.11 | 318 (7.1) | 0.14 | 347 (0.4) | 0.24 |
| Obesity | 1224 (7.4) | 2059 (8.3) | 0.03 | 288 (9.3) | 0.07 | 479 (10.6) | 0.11 | — | — |
| Dementia | 518 (3.2) | 1298 (5.2) | 0.10 | 83 (2.7) | 0.03 | 244 (5.4) | 0.11 | 138 (0.2) | 0.24 |
| **Infection predisposition** |  |  |  |  |  |  |  |  |  |
| Injection drug use proxy | 1045 (6.4) | 448 (1.8) | 0.23 | 125 (4.0) | 0.10 | 101 (2.2) | 0.20 | 242 (0.3) | 0.34 |
| Opioid use disorder | 853 (5.2) | 286 (1.1) | 0.23 | 97 (3.1) | 0.10 | 69 (1.5) | 0.20 | 135 (0.2) | 0.32 |
| Vascular catheter codes | 1296 (7.9) | 1316 (5.3) | 0.11 | 289 (9.3) | 0.05 | 501 (11.1) | 0.11 | 110 (0.1) | 0.40 |
| **Documentation intensity in ascertainment window (signed/addendum notes)** |  |  |  |  |  |  |  |  |  |
| ≥2 notes | 14 385 (87.5) | 22 174 (88.9) | 0.04 | 2754 (88.9) | 0.04 | 3980 (88.4) | 0.03 | 46 556 (56.8) | 0.73 |
| ≥10 notes | 13 934 (84.8) | 21 544 (86.4) | 0.05 | 2680 (86.5) | 0.05 | 3873 (86.0) | 0.04 | 12 557 (15.3) | 1.93 |

### Statistical Analysis

Prevalence used Wilson intervals and absolute differences used Wald intervals.^14^ Baseline characteristics are shown as counts with percentages or means with standard deviations, with standardized mean differences.

Crude odds ratios used Wald log scale intervals and Fisher exact *P* values. Finding-specific analyses used Fisher exact intervals (**eMethods 1.8** in the Supplement). The main model was a grouped binomial regression adjusted for age group, sex, race, and documentation band, with profile likelihood intervals. A modified Poisson model gave the prevalence ratio.^15^ Bands were pooled with the Mantel Haenszel estimator.^16^ Population models pooled joint age and documentation strata. Balance used standardized mean differences.^17^

The primary analysis compared SAB with *E coli* bacteremia under the locked definition. Secondary analyses used the pathology only definition against the general population, the five cohort gradient, and separate findings (**eFigures 1-2** in the Supplement). Timing was descriptive (**eTable 3** and **eFigures 3-4** in the Supplement). Sensitivity analyses included restriction to 10 or more notes, pooled and standardized estimates, other windows, and 2000 to 2025 (**eTable 4** and **eFigures 5-7** in the Supplement). Bias analysis used the Rogan Gladen correction^18^ at validated specificity, with equal sensitivity, then sensitivity 10 and 20 points lower in SAB. Post hoc analyses stratified by injection drug use and opioid use disorder and split SAB by methicillin resistance. Secondary analyses were exploratory, without multiplicity correction. Two sided *P* < .05 set the primary threshold.

The dental definition was validated in 73 patients with SAB, sampled across five signal strata and one unflagged stratum. Review used a locked rubric and was blinded to outcomes. Results are in **eTable 5** and **eFigure 8** in the Supplement. Data quality checks are in **eTable 6** in the Supplement.

## Results

### Cohort Construction and Baseline Characteristics

We identified 50 244 patients with a qualifying bacteremia diagnosis. After excluding 1 patient without a birth year and 1268 patients younger than 18 years, 48 975 adults remained: SAB in 16 435, *E coli* bacteremia in 24 942, other streptococcal sepsis in 3097, and enterococcal sepsis in 4501. The general population cohort included 81 945 adults (**Figure 1**).

**Figure 1.**
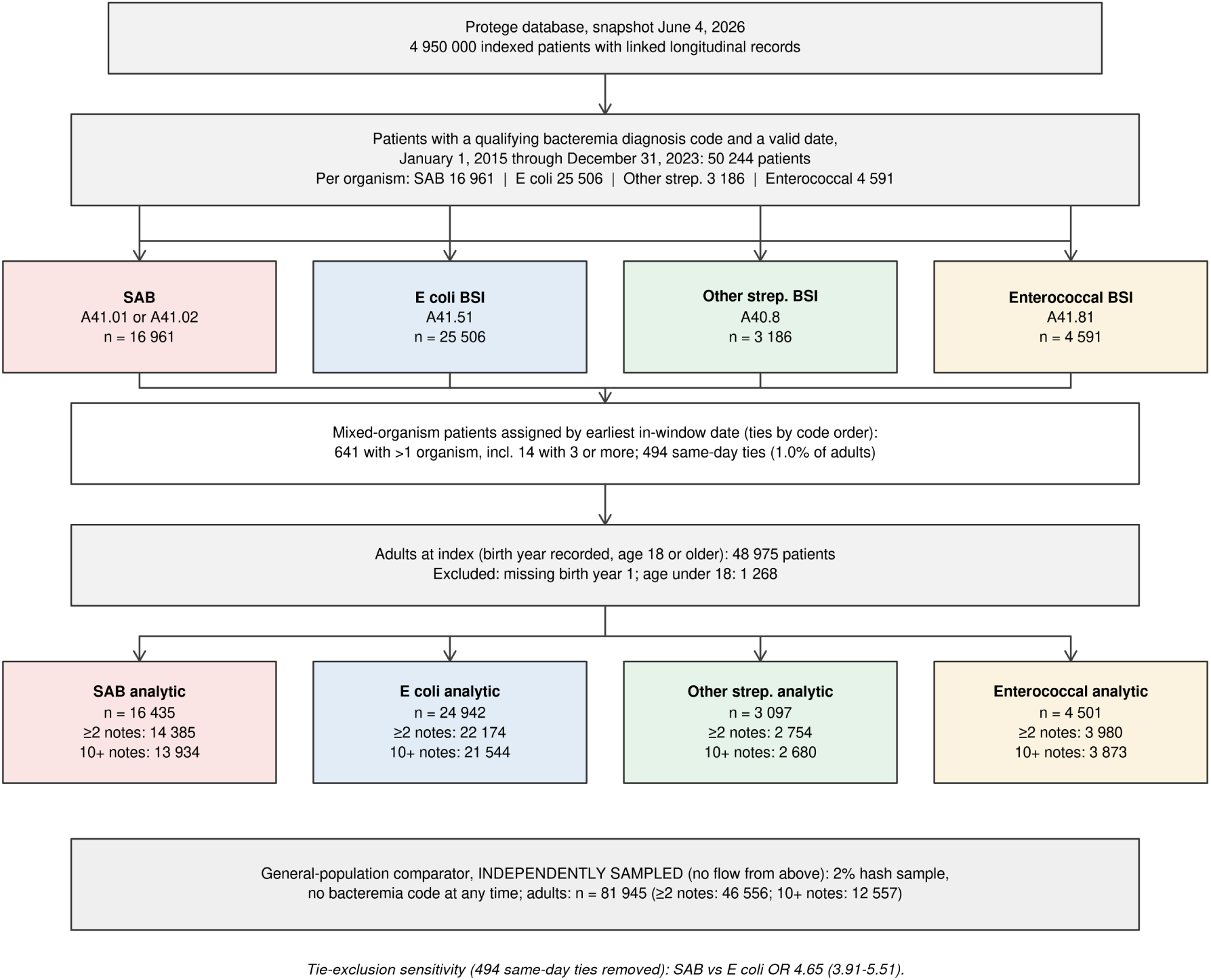
Cohort construction. Cohorts used earliest in-window diagnosis date, with same-day ties by code order. Of adults, 641 had more than 1 organism and 494 had same-day ties. The population comparator was sampled independently.

Patients with SAB were younger than those with *E coli* bacteremia (mean age, 57.9 [standard deviation, 17.2] vs 65.2 [16.7] years; standardized mean difference, 0.43) and more often male (62.1 % vs 38.3 %; standardized mean difference, 0.49) (**Table 1**). Injection drug use, opioid use disorder, end-stage kidney disease, and prior endocarditis were more common in SAB, and malignancy was less common. At least 10 notes were present for 84.8 % of SAB, 86.4 % of *E coli* bacteremia, and 15.3 % of the general population.

### Prevalence of Documented Dental Disease Across Cohorts

Pathology-only prevalence rose from 0.49 % in the general population and 0.71 % in *E coli* bacteremia to 1.91 % in enterococcal bacteremia, 3.15 % in SAB, and 7.98 % in other streptococcal bacteremia (**Figure 2**). The SAB to other streptococcal odds ratio was 0.37, with a 95% CI from 0.32 to 0.44.

**Figure 2.**
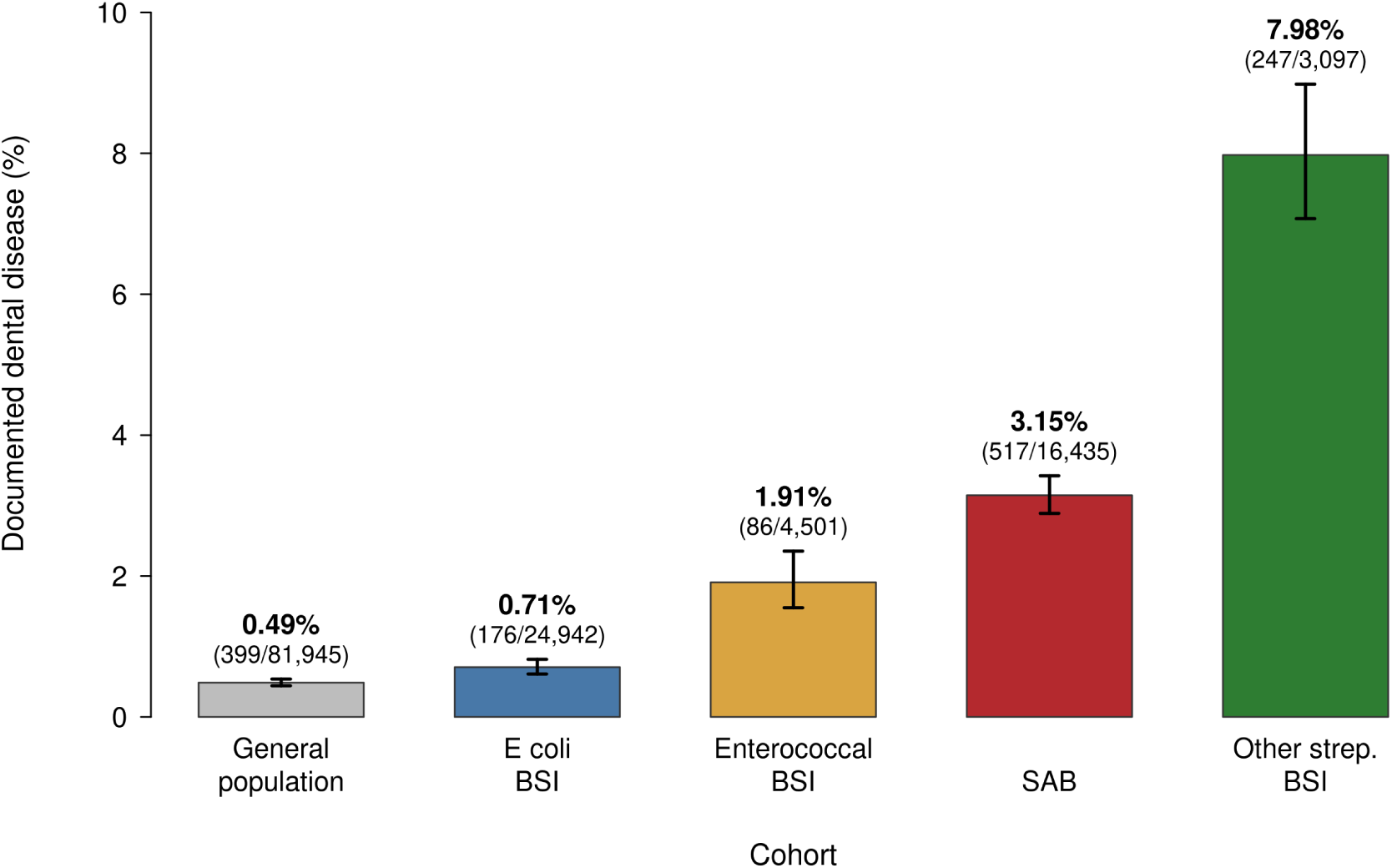
Documented dental disease across cohorts. The pathology-only definition excludes procedures. Bars show prevalence with 95% Wilson CIs. Numerators and denominators appear above each bar. No patient contributes to more than 1 bar.

### Primary Comparison: SAB vs *E coli* Bacteremia

Under the locked definition, disease was documented in 528 of 16 435 patients with SAB (3.21 %, 95% CI 2.95 to 3.49) and 179 of 24 942 patients with *E coli* bacteremia (0.72 %, 95% CI 0.62 to 0.83). The difference was 2.49 percentage points (95% CI 2.21 to 2.78). The crude odds ratio was 4.59 (95% CI 3.87 to 5.45). The adjusted odds ratio was 3.86 (95% CI 3.23 to 4.63). The prevalence ratio was 3.74 (95% CI 2.86 to 4.90) (**Table 2** and **Figure 3**).

**Table 2.** Primary and Secondary Comparisons of Documented Dental Disease.

| Comparison | Definition | SAB<br>No./total (%) | Comparator<br>No./total (%) | Difference<br>pp (95% CI) | Unadjusted<br>OR (95% CI) | Adjusted<br>OR (95% CI) |
| --- | --- | --- | --- | --- | --- | --- |
| **Primary: SAB vs *E coli* BSI** |  |  |  |  |  |  |
| Overall | Locked | 528/16 435 (3.21) | 179/24 942 (0.72) | 2.49 (2.21-2.78) | 4.59 (3.87-5.45) | 3.86 (3.23-4.63)<sup>a</sup> |
| ≥10 notes | Locked | 520/13 934 (3.73) | 178/21 544 (0.83) | 2.91 (2.57-3.24) | 4.65 (3.92-5.52) | — |
| Documentation-pooled | Locked | — | — | — | — | 4.68 (3.95-5.55)<sup>b</sup> |
| **Secondary: SAB vs general population** |  |  |  |  |  |  |
| Overall | Locked | 528/16 435 (3.21) | 408/81 945 (0.50) | 2.71 (2.44-2.99) | 6.63 (5.82-7.56) | 3.67 (3.07-4.40)<sup>c</sup> |
| Overall | Pathology-only | 517/16 435 (3.15) | 399/81 945 (0.49) | 2.66 (2.39-2.93) | 6.64 (5.82-7.57) | 3.71 (3.08-4.46)<sup>c</sup> |
| **Oral-flora-associated comparator: SAB vs other streptococcal BSI** |  |  |  |  |  |  |
| Overall | Pathology-only | 517/16 435 (3.15) | 247/3097 (7.98) | −4.83 (−5.82 to −3.84) | 0.37 (0.32-0.44) | — |
| **Contextual: SAB vs enterococcal BSI** |  |  |  |  |  |  |
| Overall | Pathology-only | 517/16 435 (3.15) | 86/4501 (1.91) | 1.24 (0.75-1.72) | 1.67 (1.32-2.10) | — |

Abbreviations: BSI, bloodstream infection; CI, confidence interval; OR, odds ratio; pp, percentage points; SAB, *Staphylococcus aureus* bacteremia; Other strep., other streptococcal (A40.8; viridans-group streptococci expected to constitute the majority). — indicates not estimated for that row. All *P* values for SAB versus *E coli* BSI and versus the general population were < .001 (2-sided Fisher exact test). The locked definition requires a dental procedure or disease-specific clinical text; the pathology-only definition excludes all procedure signals. All bacteremia comparisons use adult cohorts assigned by earliest in-window organism; the general-population comparator was sampled independently, as prespecified.
<sup>a</sup>Grouped binomial regression adjusted for age group, sex, race, and documentation band (157 populated cells, with missing sex as an explicit category; 41 377 patients; 707 events), with profile-likelihood confidence intervals. The supporting modified Poisson prevalence ratio was 3.74 (95% CI, 2.86-4.90).
<sup>b</sup>Mantel-Haenszel estimate pooled across documentation bands, with Robins-Breslow-Greenland confidence intervals.
<sup>c</sup>Mantel-Haenszel estimate pooled across joint age and documentation strata.

**Figure 3.**
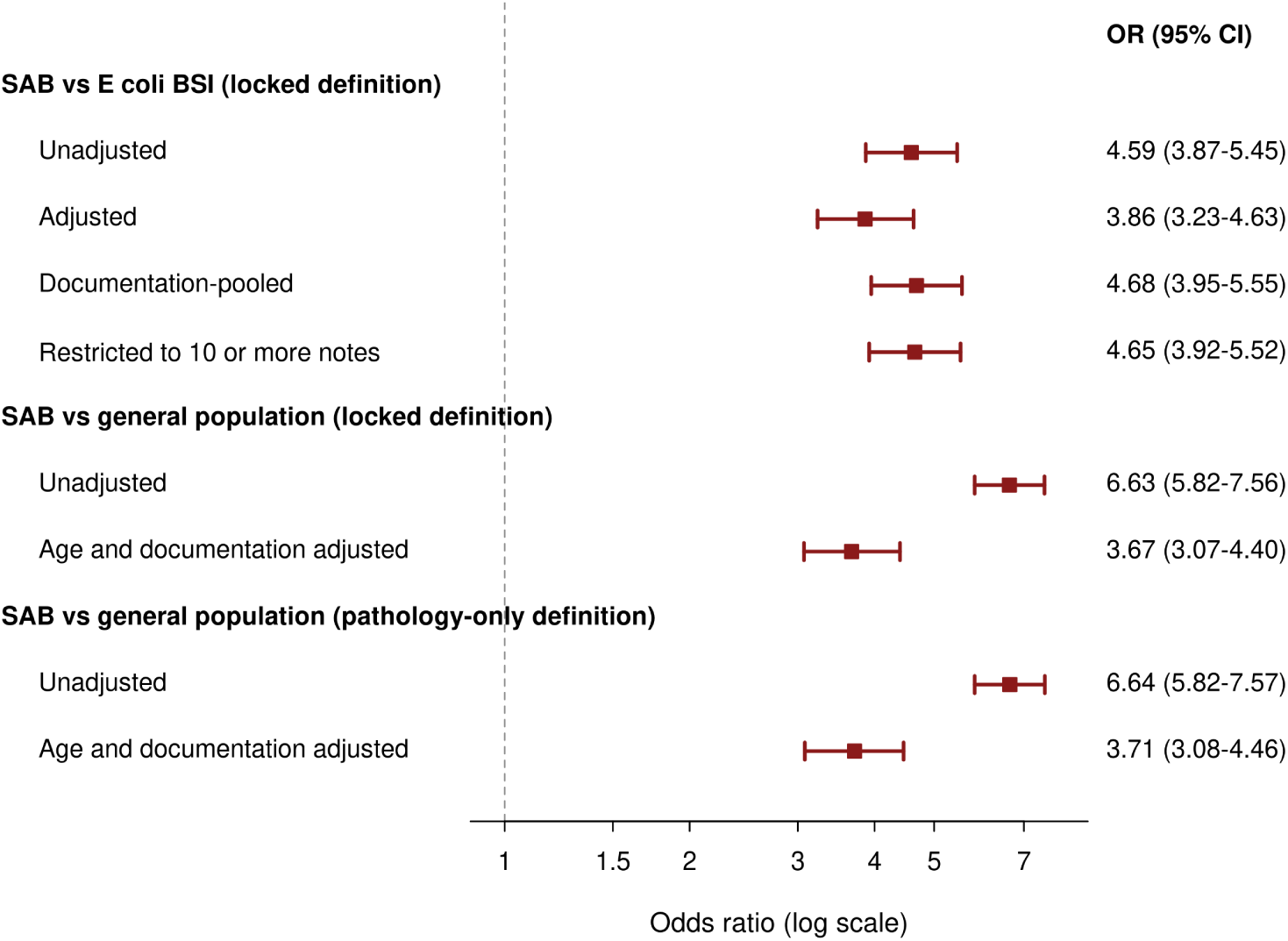
Primary and secondary comparisons in SAB. Odds ratios use a logarithmic scale. The dashed line marks the null. Adjusted and stratified specifications are in **Table 2**.

### Secondary Comparison: SAB vs the General Population

Pathology only disease was documented in 517 of 16 435 patients with SAB (3.15 %) and 399 of 81 945 population patients (0.49 %). The difference was 2.66 percentage points (95% CI 2.39 to 2.93). The crude odds ratio was 6.64 (95% CI 5.82 to 7.57). The joint age and documentation stratified odds ratio was 3.71 (95% CI 3.08 to 4.46) (**Table 2**).

### Individual Dental Findings

In patients with at least two notes, each finding was more common in SAB (**Table 3** and **Figure 4**). Dental abscess and caries gave the largest absolute gains, and retained roots, caries, and other finding gave the largest relative gains, with odds ratios from 10.14 to 14.60. Each comparison had *P* < .001. In SAB, prevalence was 5.3 % at 18 to 49 years and 1.4 % at 75 years or older. Rates were highest with prior endocarditis (9.8 %, 95% CI 7.2 to 13.2), opioid use disorder (6.2 %, 4.8 to 8.0), injection drug use (5.3 %, 4.1 to 6.8), prosthetic valve (5.0 %, 3.1 to 8.0), and native valve disease (4.8 %, 3.7 to 6.2) (**eTable 7** in the Supplement).

**Figure 4.**
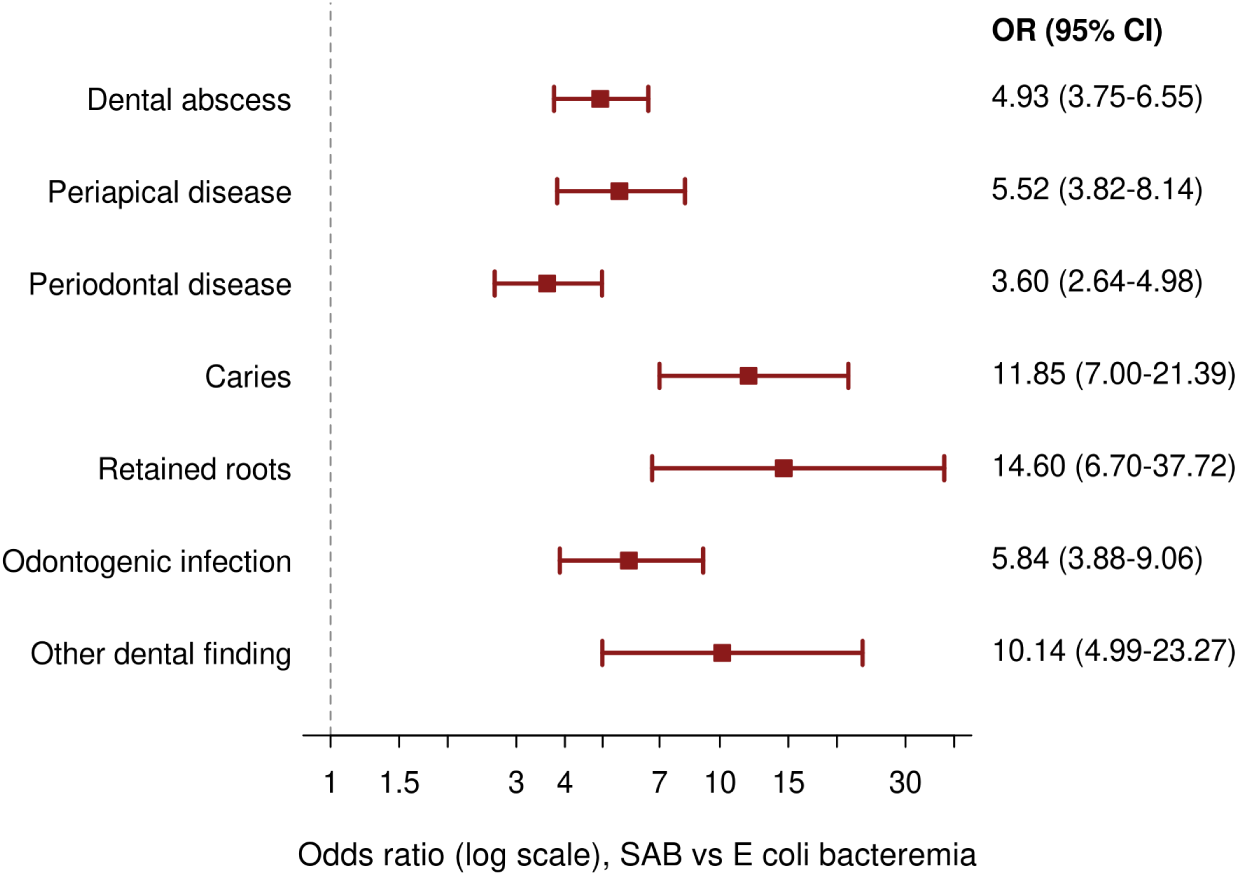
Individual dental findings, SAB vs *E coli* bacteremia. Odds ratios use a logarithmic scale. Denominators are patients with 2 or more notes. Findings overlap and do not sum to the composite outcome.

**Table 3.** Individual Dental Findings by Cohort.

| Finding | SAB<br>(n=14 385) | *E coli* BSI<br>(n=22 174) | General population<br>(n=46 556) | Other strep. BSI<br>(n=2754) | OR vs *E coli* BSI<br>(95% CI) | OR vs general population<br>(95% CI) |
| --- | --- | --- | --- | --- | --- | --- |
| Dental abscess | 221 (1.54) | 70 (0.32) | 228 (0.49) | 111 (4.03) | 4.93 (3.75-6.55) | 3.17 (2.62-3.84) |
| Periapical disease | 135 (0.94) | 38 (0.17) | 55 (0.12) | 70 (2.54) | 5.52 (3.82-8.14) | 8.01 (5.81-11.17) |
| Periodontal disease | 137 (0.95) | 59 (0.27) | 81 (0.17) | 65 (2.36) | 3.60 (2.64-4.98) | 5.52 (4.16-7.36) |
| Caries | 122 (0.85) | 16 (0.07) | 38 (0.08) | 32 (1.16) | 11.85 (7.00-21.39) | 10.47 (7.22-15.50) |
| Retained roots | 66 (0.46) | 7 (0.03) | 33 (0.07) | 28 (1.02) | 14.60 (6.70-37.72) | 6.50 (4.21-10.19) |
| Odontogenic infection | 113 (0.79) | 30 (0.14) | 40 (0.09) | 103 (3.74) | 5.84 (3.88-9.06) | 9.21 (6.36-13.56) |
| Other dental finding | 59 (0.41) | 9 (0.04) | 36 (0.08) | 16 (0.58) | 10.14 (4.99-23.27) | 5.32 (3.46-8.30) |

Values are No. (%). Abbreviations: BSI, bloodstream infection; CI, confidence interval; OR, odds ratio; SAB, *Staphylococcus aureus* bacteremia; Other strep., other streptococcal. Denominators are patients with 2 or more cleaned clinical notes in the ascertainment window (signed/addendum note-count proxy), so prevalences exceed whole-cohort values reported elsewhere. General-population finding counts use dedicated window builds joined to the fixed population index, matching the composite outcome. Findings overlap within patients and do not sum to the composite outcome. All comparisons versus *E coli* BSI and versus the general population had $P < .001$ (2-sided Fisher exact test).

### Timing Relative to Bacteremia

Among 437 patients with SAB and an assignable first-finding date, 5.7 % had the finding before index, 18.1 % on the index day, and 76.2 % (95% CI, 72.0 to 80.0) after index. The median was 3 days after index, with an interquartile range from 1 to 8 days (**eTable 3** and **eFigures 3-4** in the Supplement). The post index share was 63.9 % in *E coli* bacteremia and 80.1 % in other streptococcal bacteremia. A date was assignable for 820 of 1045 patients with disease. The rest had a first signal before the window.

### Documentation-Stratified and Sensitivity Analyses

In patients with at least 10 notes, disease was documented in 520 of 13 934 patients with SAB (3.73 %) and 178 of 21 544 patients with *E coli* bacteremia (0.83 %). The odds ratio was 4.65 (95% CI 3.92 to 5.52). Pooling across bands gave 4.68 (95% CI 3.95 to 5.55). Other specifications agreed (**eTable 4** and **eFigures 5-7** in the Supplement). Bias correction at specificity 0.995 yielded ratios from 12.46 (equal sensitivity) to 19.69 (sensitivity 20 points lower in SAB), supporting the direction but not the magnitude of the association. In 2000 to 2025, first signal disease was 2.52 % in SAB and 0.40 % in *E coli* bacteremia, with an odds ratio of 6.43 (95% CI 5.24 to 7.90).

Post hoc results agreed in direction (**eTable 8** in the Supplement), with odds ratios of 4.60 (95% CI 3.85 to 5.49) without injection drug use and 4.46 (3.74 to 5.31) without opioid use disorder. Disease was 3.12 % in methicillin susceptible SAB and 3.27 % in methicillin resistant SAB. The odds ratio was 0.96 (95% CI 0.80 to 1.14).

### Performance of the Dental Definition

Against blinded chart review, the locked definition had sensitivity of 0.55 (95% CI 0.38 to 0.71), specificity of 1.00, positive predictive value of 0.88, and negative predictive value of 0.97. Full results are in **eTable 5** and **eFigure 8** in the Supplement.

## Discussion

Dental disease was documented in 3.21% of patients with SAB and 0.72% of patients with *E coli* bacteremia. The adjusted odds ratio was 3.86 (95% CI, 3.23 to 4.63). Disease was also more common in SAB than in the general population after adjustment, with an odds ratio of 3.71 (95% CI, 3.08 to 4.46), and the excess persisted in patients with at least 10 notes. Among patients with SAB and an assignable date, 76.2% had the first finding recorded after the index date.

Earlier work on oral portals focused mainly on viridans group streptococcal infection.^8,9^ A systematic study of infective endocarditis found an oral or dental portal in a minority of patients, but it included no nonoral bacteremia comparison.^10^ Reviews of SAB stress source control, yet they have not quantified dental disease.^5^ SAB sat between enterococcal and other streptococcal bacteremia. The SAB versus enterococcal odds ratio was 1.67 (95% CI, 1.32 to 2.10), compared with a crude SAB versus *E coli* odds ratio of 4.59 (95% CI, 3.87 to 5.45). Enterococcal bacteremia also prompts endocarditis evaluation, so that contrast may be less affected by how hard clinicians search the mouth. Its smaller size suggests that part of the *E coli* contrast reflects detection during workup.

Several pathways may explain this pattern. Oral activity and dental procedures can introduce oral organisms into the bloodstream,^9,19^ and *S aureus* colonizes the mouth in health and disease.^20^ The large relative differences for caries and retained roots suggest a higher burden of chronic dental disease in SAB. Within SAB, disease was most common in younger patients and in patients with prior endocarditis, opioid use disorder, injection drug use, or valve disease. Rates were lower in diabetes, chronic kidney disease, malignancy, and heart failure. Targeted detection remains a strong alternative explanation, because high-risk patients receive closer endocarditis evaluation and dental review may precede valve surgery. The excess was not confined to those groups. Periodontal disease remains common in US adults.^21^

Timing urges caution. Most findings were first recorded after the index date, when bacteremia had already triggered a source search, so these data cannot distinguish disease that contributed to infection from disease found during evaluation.

A focused oral history and examination are already part of standard SAB and endocarditis evaluation. These results support oral assessment when no source is apparent, dentition is poor, endocarditis is suspected, or a prosthetic valve is present. Abnormal findings warrant dental or oral surgery consultation, without replacing evaluation for endocarditis, metastatic infection, or device infection.^5,6^ This study did not test dental treatment or imaging. An absent dental record should not reassure, because sensitivity was 0.55. Prospective study should use a standardized oral examination.

### Limitations

This study measured documented disease, not disease on standardized oral examination, so detection bias is central. SAB prompts a more intensive search than *E coli* bacteremia. Documented prevalence mixes disease present before bacteremia with disease found during workup. Sensitivity was low. The bias analysis supported direction, but not a precise corrected magnitude. Cohorts were defined by prespecified ICD-10-CM codes, as is standard in large electronic health record studies,^13^ so misclassification remains possible. Procedure data captured dental surgery poorly, so note text was needed. The population cohort differed in age and note volume. Adjustment reduced its odds ratio from 6.64 to 3.71, but residual ascertainment bias likely remains. No fully workup matched comparator was available. Actual workup received was not measured. Dental consultation was not a covariate, because it lies on the workup pathway. Direct workup measures and a negative control outcome were unavailable. Validation also covered SAB only, with one reviewer and no second reader, so differential misclassification between cohorts could not be excluded. Timing was first documentation, not onset, so a post index record does not show that disease began after bacteremia. Socioeconomic position, smoking, and dental care access were unavailable, and generalizability is uncertain. The design cannot establish the mouth as the portal of entry.

## Conclusions

Documented dental disease was more common in SAB than in *E coli* bacteremia or the general population, but most findings were recorded only after bacteremia was recognized. These data do not establish the mouth as the source of SAB. Prospective studies with standardized oral examination are needed.

*Concept and design:* Ssentongo.

*Acquisition, analysis, or interpretation of data:* Ssentongo.

*Drafting of the manuscript:* Ssentongo.

*Critical review of the manuscript for important intellectual content:* Ssentongo, Peacock, Durieux, Colovos.

*Statistical analysis:* Ssentongo, Durieux.

*Supervision:* Ssentongo.

## Supporting information

Supplemental Online Content

## Data Availability

The Protege data are not publicly available and access is governed by the data owner. The study protocol, statistical analysis plan, code lists and aggregate results are available from the Corresponding author on reasonable request.

## Data Sharing Statement

The Protégé data are not publicly available and access is governed by the data owner. The study protocol, statistical analysis plan, code lists and aggregate results are available from the Corresponding author on reasonable request.

## Additional Contributions

We thank Myriam Amsallem, MD, for critical review of and comments on the manuscript.

## Ethics and Reporting

Ethics approval and consent to participate: This study was conducted in accordance with guidelines of the Declaration of Helsinki, Belmont report, and US Common Rule. Before delivery data was de-identified through expert determination. An external review by Bluebonnet Ethical Review determined that this activity is appropriately classified as Not Human Subjects Research (NHSR) and that IRB review is not required. Consent for publication: Not applicable.

## AI Disclosure

This study was supported through using an internal Meta scaffold around Muse Spark 1.3 and an internal version of the next generation of Muse models, which assisted the authors in all stages of the study. Advisory agent critiques and coauthor review informed the scientific revisions. The authors adjudicated all recommendations. No AI system is an author. Authors remain responsible for the accuracy of all methods, results, references, and declarations.

