## Supplemental Online Content for "Documented Dental Disease in Patients With *Staphylococcus aureus* Bacteremia"

Paddy Ssentongo, MD, PhD, W. Frank Peacock, MD, FACEP, FACC, FESC, Jared C. Durieux, PhD, MS, MDS, MPH, Christos Colovos, MD, PhD

Meta Platforms, Inc., Menlo Park, California, USA. Department of Medicine, Division of Infectious Diseases, Penn State Health Milton S. Hershey Medical Center, Hershey, Pennsylvania, USA. Department of Public Health Sciences, Penn State College of Medicine, Hershey, Pennsylvania, USA. Henry JN Taub Department of Emergency Medicine, Baylor College of Medicine, Houston, Texas, USA. Evidence Architecture, Stow, Ohio, USA

2026

All authors are consultants or contingent workers for Meta Platforms, Inc.

**Table of Contents**

**eMethods 1.** Data source, cohort construction, exposure definition, and analytic specification
**eTable 1.** Complete phenotype definitions, code lists, and text patterns
**eTable 2.** Baseline characteristics of the enterococcal bloodstream infection cohort
**eTable 3.** Timing of the first documented dental finding relative to bacteremia
**eTable 4.** Prespecified sensitivity, standardization, and quantitative bias analyses
**eTable 5.** Performance of the dental definitions against blinded chart review
**eTable 6.** Data-quality and measurement checks
**eTable 7.** Individual dental findings and pathology-only dental disease in SAB by baseline characteristic
**eTable 8.** Stratified and organism-subtype sensitivity analyses
**eFigure 1.** Prevalence of individual dental findings by cohort
**eFigure 2.** Finding-specific odds ratios, SAB vs the general population
**eFigure 3.** Timing of the first dental finding relative to bacteremia, by cohort
**eFigure 4.** Daily distribution of first dental findings in SAB
**eFigure 5.** Documented dental disease by documentation intensity
**eFigure 6.** Age-stratified prevalence, SAB vs the general population
**eFigure 7.** Prespecified sensitivity and bias analyses
**eFigure 8.** Performance of the dental definition against blinded chart review

### eMethods 1. Data Source, Cohort Construction, Exposure Definition, and Analytic Specification

#### 1.1 Data source

This retrospective study used the Protégé research database (June 2026 release), a de-identified, longitudinal database that links electronic health records (EHR) with open medical and pharmacy claims. Protégé draws on routine-care EHR data from a network of 39 medium-to-large US health systems. Subsets of patients are also linked to radiology data and to dated records of death. The release contains 4 950 000 patients residing in all 50 US states and the District of Columbia, with 8.6 billion clinical events and 775 million clinical notes. The median record span, from first to last dated clinical event, is 11.4 years (interquartile range, 7.0 to 18.8), and dated events fall predominantly between 2008 and 2024. Structured EHR data include demographics, encounters in ambulatory, telehealth, emergency, inpatient, and observation settings, diagnoses (predominantly ICD-10-CM), procedures (CPT, HCPCS, ICD-10-PCS, and local codes), laboratory results, vital signs, medication orders and fills, and surgical cases. Clinical notes retain their original text. Claims data include medical claim lines and headers with diagnosis and procedure codes and payer type, and pharmacy claims with National Drug Codes, days supply, and dispensing status, and 99.4% of patients have linked medical or pharmacy claims. For this study, we used data from January 1, 2015, to December 31, 2023 for the primary analyses. We used the snapshot dated June 4, 2026. That snapshot had 8 583 623 859 event records and 775 419 297 clinical notes. All queries ran inside the governed environment on Presto SQL. No record level data left that environment. No figure and no table in this study carries a patient identifier.

#### 1.1a Database construction

The database was built from aggregator patients with at least five clinical notes and records linkable to open claims. Patients linked to radiology data were included preferentially. The remaining sample was enriched for diagnostic diversity by preferentially oversampling low-prevalence clinical phenotypes within the ICD-10 hierarchy. The database is therefore not a probability sample of the US population or of the contributing health systems' patients.

#### 1.1b Scope and de-identification

The data capture care documented by contributing health systems and services billed in open claims. Insurance enrollment periods are not available, so periods without recorded care cannot be distinguished from care received elsewhere. Cost information is limited to billed charges, mortality is all-cause, and facility identifiers are not included. Data were de-identified before delivery under the HIPAA Expert Determination method (45 CFR 164.514(b)(1)), birth dates are reduced to the year, ages of 89 years and older are top-coded, residence is reported as state and 3-digit ZIP code, and names of people and places in clinical text are replaced with synthetic surrogates. Before delivery data was de-identified through expert determination. An external review by Bluebonnet Ethical Review determined that this activity is appropriately classified as Not Human Subjects Research (NHSR) and that IRB review is not required.

##### 1.2 Bacteremia cohort construction

We built the bacteremia cohorts from diagnosis records that carried an electronic health record source and a valid date. The qualifying codes were A41.01 and A41.02 for SAB, A41.51 for *Escherichia coli*, A40.8 for other streptococcal sepsis, and A41.81 for enterococcal sepsis. The A40.8 cohort is a code defined other streptococcal sepsis group. It is not a single species group. Viridans group streptococci likely form most of that group on clinical grounds, but the code itself does not prove species. For each patient, the index date was the earliest qualifying date for an organism inside the calendar window. Patients were eligible when they had a recorded birth year and were adults at index, unless a table states a different rule.

Each patient was assigned to one organism only. The base build had 50 244 patients with a qualifying bacteremia diagnosis. Assignment by the earliest in-window date gave SAB 16 961, *E coli* 25 506, other streptococcal sepsis 3186, and enterococcal sepsis 4591. One patient had no recorded birth year. A further 1268 patients were under 18 years old. The adult cohorts were SAB 16 435, *E coli* 24 942, other streptococcal sepsis 3097, and enterococcal sepsis 4501. The adult total was 48 975. In the full window, 641 patients carried more than one organism code. Each such patient was counted once, under the earliest date. On the earliest date, 494 patients had a tie between organisms, which is 1.0% of adults. Ties were resolved by code order, with the lowest ICD-10-CM code assigned.

Before assignment, the pairwise overlaps were as follows. SAB and *E coli* overlapped in 130 patients. SAB and other streptococcal sepsis overlapped in 51 patients. SAB and enterococcal sepsis overlapped in 123 patients. *E coli* and other streptococcal sepsis overlapped in 63 patients. *E coli* and enterococcal sepsis overlapped in 265 patients. Other streptococcal sepsis and enterococcal sepsis overlapped in 37 patients. A total of 14 patients carried three or more organism codes. When we removed the tied patients in a sensitivity analysis, the unadjusted odds ratio was 4.65, with a 95% CI from 3.91 to 5.51.

##### 1.3 Calendar window

The primary window was January 1, 2015, through December 31, 2023. Dates are shifted within each patient by the vendor. Intervals inside a record remain exact. Calendar year labels should be read as near years, not exact years. We locked the window once we had reviewed capture by year. SAB counts were stable from 2015 through 2023. They then fell to about 51% of the prior year level in the last complete year. That last complete year had 1312 SAB patients, compared with 2566 before it, under a first qualifying index rule. The counts also declined inside that year, which fits incomplete accrual more closely than a true change in disease rate. The final partial year had fewer than 25 patients and no dental text findings. In the final two years, about 16% of index dates had less than a full 30 day ascertainment window. The share ran from 15.8 to 16.3% across index rules. If partial windows had been admitted, later years would have had systematically shorter follow-up. The full 2000 to 2025 record set was retained for sensitivity analysis only.

##### 1.4 General-population comparator

We drew the general population comparator as a 2% deterministic hash sample. The hash used the tokenized patient identifier with the seed value 20260920, through the Presto expression recorded in each cohort build manifest. The rule makes the sample reproducible. The sample does not depend on the outcome. To enter the sample, a patient needed no qualifying bacteremia code at any time. The patient also needed a recorded birth year and a first observed health care contact inside the calendar window. That first contact was the pseudo index. The build held 96 433 patients before the adult age filter. It held 81 945 adults for analysis. The pseudo index is an encounter based stand-in for first health care contact.

Outcome work and baseline work use the same adult denominator, 81 945. At sampling, all qualifying bacteremia codes were excluded without regard to date.

#### 1.5 Exposure and outcome definition

The ascertainment window was the same for all cohorts. It ran from index day minus 7 through index day plus 30. Two definitions were used.

The locked definition required one of two signals. The first signal was a qualifying dental procedure code. The second signal was a qualifying disease specific text finding. We used this definition for the primary SAB to *E coli* comparison and for the locked definition population comparison.

The pathology only definition required a qualifying disease specific text finding. It removed every procedure signal, including extraction. We used this definition for the cross cohort prevalence gradient, for the control comparisons, and for the primary population comparison. The reason for the separate definition is age linked procedure use. Dental procedures in a general population sample often reflect planned third molar removal in young adults. That pattern differs from procedures carried out during an acute bacteremia illness. A single procedure inclusive definition would therefore compare unlike exposures across cohorts.

Text matching was insensitive to case. Each pattern required an explicit word boundary. Text had to appear in a signed note or an addendum note. The full patterns are in eTable 1.

#### 1.6 Prespecified exclusions and code-collision audit

We audited candidate codes before we estimated any comparison. We removed code groups that matched nondental care. The obstetric and larynx groups were `10D*` and `0C5*`. The imaging code `IMG8005` matched dental terms by substring in a skeletal survey. The imaging code `IMG850` matched an odontoid view. The plain code `2313` matched an alpha-fetoprotein laboratory row. After those removals, the extraction flagged count fell from 61 to 52 patients, and the any signal count fell from 145 to 136 patients, in the first narrow test window. We barred two candidate rules from use as exposures. The broad any dental mention rule found 11 447 patients, and note templates made it unreliable. Dental note type alone had almost no capture inside the window, so it added no signal.

#### 1.7 Covariates and documentation intensity

We read comorbidities from diagnosis codes in the 365 days before index, without the index day itself. We measured documentation intensity as the count of cleaned clinical notes inside the ascertainment window. We then grouped that count as 0 to 1 notes, 2 to 9 notes, or 10 or more notes. We handled this count as a confounder. The outcome exists in the data only when a patient is examined and the finding is written down. Note volume therefore affects both the chance of capture and the groups being compared.

#### 1.8 Statistical specification

Let $x$ denote patients meeting the dental definition and let $n$ denote the cohort denominator. Prevalence is $\hat{p} = x/n$. Its Wilson score interval is

$$\left(\hat{p} + \frac{z^2}{2n} \pm z\sqrt{\frac{\hat{p}(1-\hat{p})}{n} + \frac{z^2}{4n^2}}\right) \Big/ \left(1 + \frac{z^2}{n}\right),$$

where $z = 1.96$. The absolute difference in prevalence uses the Wald interval

$$(\hat{p}_1 - \hat{p}_2) \pm z\sqrt{\frac{\hat{p}_1(1-\hat{p}_1)}{n_1} + \frac{\hat{p}_2(1-\hat{p}_2)}{n_2}}.$$

The crude odds ratio is $\mathrm{OR} = ad/(bc)$. In that expression, $a$ and $b$ are exposed patients with and without the outcome. The terms $c$ and $d$ are the matching comparator counts. The standard error is $\mathrm{SE}\{\ln \mathrm{OR}\} = \sqrt{1/a + 1/b + 1/c + 1/d}$. We report two sided Fisher exact $P$ values.

The prespecified adjusted estimate came from a grouped binomial model over covariate cells. The model is

$$\mathrm{logit}(\pi_i) = \beta_0 + \beta_1 X_i + \boldsymbol{\gamma}'\mathbf{Z}_i.$$

The fit uses maximum likelihood over the cell counts. The cell log likelihood is $\ell = \sum_i [y_i \ln \pi_i + (m_i - y_i)\ln(1-\pi_i)]$, where $m_i$ is the patient count in cell $i$ and $y_i$ is the event count in that cell. Cells crossed four age groups, 18 to 49, 50 to 64, 65 to 74, and 75 years or older. Cells also crossed sex, male, female, or missing as its own category, and race, White, Black, or other or unknown, and three documentation bands. The model had 157 populated cells. It used 41 377 patients and 707 events. Intervals for the grouped model are profile likelihood intervals. They come from inversion of the likelihood ratio test at the 95% level.

The prevalence ratio came from a modified Poisson model. The model uses a log link and robust variance. It is

$$\ln(\pi_i) = \alpha + \theta X_i + \boldsymbol{\delta}'\mathbf{Z}_i, \qquad \mathrm{Var}(\hat{\boldsymbol{\beta}}) = \mathbf{H}^{-1}\mathbf{M}\mathbf{H}^{-1},$$

where $\mathbf{H}$ is the Hessian at the fitted values and $\mathbf{M} = \sum_i \mathbf{u}_i\mathbf{u}_i'$ is the meat matrix from cell score contributions $\mathbf{u}_i$.

Pooled strata used the Mantel Haenszel estimator. That estimator is $\mathrm{OR}_{\mathrm{MH}} = \sum_k a_k d_k/N_k \big/ \sum_k b_k c_k/N_k$. Its interval uses the Robins Breslow Greenland variance for $\ln \mathrm{OR}_{\mathrm{MH}}$. We measured baseline balance with the standardized mean difference. For a binary trait, that measure is $d = (p_1 - p_2)/\sqrt{[p_1(1-p_1) + p_2(1-p_2)]/2}$. Direct standardization uses the combined cohort distribution of the standard variable. It then applies that distribution to each cohort rate by stratum.

Bias analysis used the Rogan Gladen correction. The correction is $p_{\mathrm{corr}} = (p_{\mathrm{obs}} + \mathrm{Sp} - 1)/(\mathrm{Se} + \mathrm{Sp} - 1)$. We used it in each cohort on its own. We held specificity at 0.995. We set sensitivity equal across cohorts, then 10 points lower in SAB, then 20 points lower in SAB. Interval width was carried forward by the delta method.

Analysis took place in September 2026. Point estimation used Python 3 standard library code. The code was checked against closed form results. Figures were rendered in R version 4.1.2.

#### 1.9 Analysis hierarchy

The primary analysis compared SAB with *E coli* bacteremia under the locked definition. The cohorts were disjoint. Secondary analyses compared SAB with the general population under the pathology only definition. They also covered the five cohort prevalence gradient and each dental finding on its own. Prespecified sensitivity analyses were restriction to the band with 10 or more notes, pooling across documentation bands, joint stratification, direct standardization, alternate windows of 0 to plus 14, 0 to plus 30, and minus 7 to plus 14 days, the full 2000 to 2025 calendar span, and quantitative bias analysis. The 10 or more notes restriction was the main robustness check. Timing analyses were descriptive. No results outside this list are reported. Secondary and sensitivity analyses were read as exploratory. No adjustment for multiple comparisons was applied.

##### 1.10 Validation design

We drew a stratified random sample of 73 patients with SAB from the full frame. The design used five signal strata and one unflagged stratum. The strata were procedure flagged, with 11 of 11 sampled, dental or oral source text, with 12 of 1706 sampled, dental clearance or valve surgery text, with 12 of 840 sampled, broad dental finding terms, with 12 of 355 sampled, multiple signals, with 14 of 456 sampled, and no structured dental signal, with 12 of 16 601 sampled. Sampling fractions therefore ran from 1.0 for the procedure flagged stratum down to 0.0007 for the unflagged stratum. One reviewer read each chart against a rubric that was locked before review. During adjudication, the reviewer did not see cohort comparison results or any downstream clinical outcome. We corrected the operating characteristics with inverse sampling weights. Population weighted estimates have an effective sample size near 17. Claims about unflagged patients therefore rest on 12 charts, and they should be read with that limit.

##### 1.11 Use of model assistance

Model assistance for this study is disclosed in the Article Information section of the main manuscript.

#### Supplementary Tables

**eTable 1. Complete Phenotype Definitions, Code Lists, and Text Patterns**

| Element | Definition |
|---|---|
| **Cohort-defining codes (ICD-10-CM, EHR source, valid date)** | |
| SAB | A41.01, A41.02 |
| *E coli* BSI | A41.51 |
| Other streptococcal BSI | A40.8 (code-defined other streptococcal sepsis, viridans-group streptococci expected majority) |
| Enterococcal BSI | A41.81 |
| General population | No qualifying bacteremia code at any time, 2% deterministic hash sample, recorded birth year, first health care contact within the calendar window serves as pseudo-index |
| **Ascertainment window** | |
| Primary | Index day −7 through index day +30 |
| Sensitivity windows | 0 to +14, 0 to +30, −7 to +14 |
| **Dental disease: text patterns (case-insensitive, word-boundary matched)** | |
| Periapical disease | periapical lucency, periapical radiolucency, periapical infection, periapical abscess |
| Dental or odontogenic abscess | dental abscess, tooth abscess, odontogenic abscess |
| Advanced caries | advanced caries, gross caries, multiple caries, severe caries, advanced decay, gross decay, multiple decay, severe decay, carious lesion(s) |
| Retained roots | retained root(s), root tip(s), root fragment(s), root remains |
| Periodontal disease | periodontal disease, periodontitis |
| **Dental disease: composite rules** | |
| Locked definition | Qualifying dental procedure OR any qualifying text pattern above |
| Dental procedures (ICD-10-PCS) | 0CDWXZ0, 0CDWXZ1, 0CDWXZ2, 0CDXXZ0, 0CDXXZ1, 0CDXXZ2, 0CTW0Z0, 0CTWXZ0 |
| Dental procedures (CDT, via HCPCS) | D7140, D7210, D7220, D7230, D7240, D7241, D7250, D7251 |
| Pathology-only definition | Any qualifying text pattern above, all procedure signals excluded, including extraction |
| **Prohibited rules (not used as exposures)** | |
| Broad dental mention | Any dental-related term anywhere in the note, identified 11 447 patients, contaminated by templated oral-examination language, inadequate positive predictive value |
| Dental note type alone | Essentially no capture within the ascertainment window |
| **Excluded code families (audited collisions, removed before analysis)** | |
| `10D*` | Obstetric extraction family (eg, 10D17ZZ) |
| `0C5*` | Larynx and pharynx procedure family |
| `IMG8005` | Skeletal survey imaging, matched dental identifiers on substring |
| `IMG850` | Odontoid view imaging |
| `2313` (undotted) | Collided with alpha-fetoprotein laboratory rows |
| **Covariates (diagnosis codes, 365 days before index, index day excluded)** | |
| Diabetes | E08, E09, E10, E11, E13 |
| Chronic kidney disease | N18 |

*eTable 1 (continued)*

| Element | Definition |
|---|---|
| End-stage kidney disease or dialysis | N18.6, Z99.2, Z49 |
| Liver disease | K70, K71.7, K74, K76 |
| COPD | J41, J42, J43, J44 |
| Malignancy | C (any) |
| HIV or immunocompromise | B20, D80-D84, Z94 |
| Obesity | E66 |
| Heart failure | I50 |
| CAD or prior MI | I20-I25 |
| Cerebrovascular disease | I60-I63, I65-I69 |
| Peripheral vascular disease | I70-I73 |
| Native valve disease | I34-I37 |
| Prosthetic valve | Z95.2, Z95.3, Z95.4 |
| Cardiac device | Z95.0, Z95.810, Z95.811 |
| Prior infective endocarditis | I33 |
| Dementia | F00-F03, G30 |
| Opioid use disorder | F11 |
| Injection drug use proxy | F11, B17, B18.2 |
| Vascular catheter codes | Z45.2, T82.7, Z95.828 |
| **Documentation intensity** | |
| Definition | Count of cleaned clinical notes within the ascertainment window |
| Bands | 0-1, 2-9, ≥10 notes |

**eTable 2. Baseline Characteristics of the Enterococcal Bloodstream Infection Cohort**

| Characteristic | Value | Characteristic | Value |
|---|---|---|---|
| Total, No. | 4501 | Age, mean (SD), y | 66.8 (14.9) |
| Male | 2785 (61.9) | White | 3206 (71.2) |
| Black | 647 (14.4) | Asian | 229 (5.1) |
| Other or unknown race | 419 (9.3) | Diabetes | 1287 (28.6) |
| Chronic kidney disease | 876 (19.5) | End-stage kidney disease | 227 (5.0) |
| Liver disease | 530 (11.8) | COPD | 575 (12.8) |
| Malignancy | 1114 (24.8) | HIV or immunocompromise | 318 (7.1) |
| Obesity | 479 (10.6) | Dementia | 244 (5.4) |
| Heart failure | 914 (20.3) | CAD or prior MI | 1069 (23.8) |
| Cerebrovascular disease | 566 (12.6) | Peripheral vascular disease | 644 (14.3) |
| Native valve disease | 571 (12.7) | Prosthetic valve | 232 (5.2) |
| Cardiac device | 292 (6.5) | Prior infective endocarditis | 89 (2.0) |
| Injection drug use proxy | 101 (2.2) | Opioid use disorder | 69 (1.5) |
| Vascular catheter codes | 501 (11.1) | ≥2 notes | 3980 (88.4) |
| ≥10 notes | 3873 (86.0) | — | — |

Values are No. (%) unless otherwise indicated. Abbreviations: CAD, coronary artery disease, COPD, chronic obstructive pulmonary disease, HIV, human immunodeficiency virus, MI, myocardial infarction, SD, standard deviation. Comorbidities were ascertained from diagnosis codes during the 365 days before index. The cohort comprises adults assigned by earliest in-window organism (n=4501), matching the enterococcal column of **Table 1** in the main article, enterococcal bacteremia is reported as a contextual comparator in **Figure 2**.

#### eTable 3. Timing of the First Documented Dental Finding Relative to Bacteremia

| Interval | SAB (n=437) | *E coli* BSI (n=108) | Other strep. BSI (n=211) | Definition |
|---|---|---|---|---|
| Before index (days −7 to −1) | 25 (5.7) | 16 (14.8) | 11 (5.2) | Documented before bacteremia |
| Index day | 79 (18.1) | 23 (21.3) | 31 (14.7) | Same calendar date |
| After index (days +1 to +30) | 333 (76.2) | 69 (63.9) | 169 (80.1) | Newly documented |
| Days 1-3 | 151 (34.6) | 28 (25.9) | 80 (37.9) | |
| Days 4-7 | 71 (16.2) | 21 (19.4) | 52 (24.6) | |
| Days 8-14 | 67 (15.3) | 8 (7.4) | 21 (10.0) | |
| Day ≥15 | 44 (10.1) | 12 (11.1) | 16 (7.6) | |
| Index day or later | 412 (94.3) | 92 (85.2) | 200 (94.8) | 95% CI for SAB, 91.7-96.1 |
| Median day (IQR) | +3 (1 to +8) | +1 (0 to +6) | +3 (1 to +5) | Order statistics |

Values are No. (%) of patients with an ascertainable first-finding date (820 of 1045 patients with documented disease, 78.5%, remainder had first-ever signals predating the window). Abbreviations: BSI, bloodstream infection, CI, confidence interval, IQR, interquartile range, SAB, *Staphylococcus aureus* bacteremia, Other strep., other streptococcal. The date of first documentation is not the date of disease onset, chronic findings such as caries and retained roots necessarily predate the note that records them.

#### eTable 4. Prespecified Sensitivity, Standardization, and Quantitative Bias Analyses

| Analysis | Comparison | Estimate (95% CI) | Specification |
|---|---|---|---|
| **Primary analysis and adjustment** | | | |
| Unadjusted OR | SAB vs *E coli* | 4.59 (3.87-5.45) | Locked definition, earliest-organism cohorts |
| Adjusted OR | SAB vs *E coli* | 3.86 (3.23-4.63) | Age, sex (missing as an explicit category), race, documentation band, 157 cells, N=41 377 |
| Adjusted prevalence ratio | SAB vs *E coli* | 3.74 (2.86-4.90) | Modified Poisson, robust variance |
| **Documentation** | | | |
| Documentation-pooled OR | SAB vs *E coli* | 4.68 (3.95-5.55) | Mantel-Haenszel across 3 bands |
| Restricted to ≥10 notes | SAB vs *E coli* | 4.65 (3.92-5.52) | Common documentation band |
| Documentation-pooled OR | SAB vs population | 3.24 (2.71-3.88) | Mantel-Haenszel across 3 bands, locked definition |
| **Standardization** | | | |
| Band-standardized prevalence | SAB vs *E coli* | 3.25% vs 0.71% | Combined cohort band distribution |
| Age-standardized prevalence | SAB vs *E coli* | 2.82% vs 0.75% | Combined cohort age distribution |
| **Population comparison** | | | |
| Joint age and documentation OR | SAB vs population | 3.67 (3.07-4.40) | Locked definition |
| Joint age and documentation OR | SAB vs population | 3.71 (3.08-4.46) | Pathology-only definition |
| **Calendar period** | | | |
| Same-day ties excluded | SAB vs *E coli* | 4.65 (3.91-5.51) | Unadjusted OR, 494 tied adults removed |
| Full 2000-2025 period | SAB vs *E coli* | 6.43 (5.24-7.90) | 473/18 783 vs 114/28 498, first-signal dates |
| In-era 2015-2023 | SAB vs *E coli* | 6.17 (4.99-7.64) | 425/16 313 vs 107/24 791, first-signal dates |
| **Ascertainment windows (first-signal dates, undercounts, *E coli*-differential)** | | | |
| Window 0 to +14 days | SAB vs *E coli* | 7.12 (5.57-9.19) | Pattern-stable, absolute values biased up |
| Window 0 to +30 days | SAB vs *E coli* | 6.95 (5.52-8.81) | Pattern-stable, absolute values biased up |
| Window −7 to +14 days | SAB vs *E coli* | 6.34 (5.06-8.02) | Pattern-stable, absolute values biased up |
| **Quantitative bias analysis (specificity fixed at 0.995)** | | | |
| Equal sensitivity | SAB vs *E coli* | 12.46 (7.62-20.37) | Rogan-Gladen corrected PR |
| Sensitivity 10 points lower in SAB | SAB vs *E coli* | 15.26 (9.34-24.95) | Rogan-Gladen corrected PR |
| Sensitivity 20 points lower in SAB | SAB vs *E coli* | 19.69 (12.04-32.18) | Rogan-Gladen corrected PR |

Abbreviations: CI, confidence interval, OR, odds ratio, PR, prevalence ratio, SAB, *Staphylococcus aureus* bacteremia. Mantel-Haenszel estimates use Robins-Breslow-Greenland confidence intervals. Window-based sensitivity analyses use study-wide first-signal dates with local windowing (78.5% capture, disproportionate *E coli* undercount), so their absolute odds ratios are biased upward and only the stable elevation pattern is interpretable. The bias analysis corrects for imperfect sensitivity at fixed high specificity: because the comparator prevalence lies near the false-positive rate, correction inflates rather than attenuates the ratio, confirming direction rather than magnitude.

**eTable 5. Performance of the Dental Definitions Against Blinded Chart Review**

| Measure | Dental evaluation rule (n=69 evaluable) | Dental pathology rule (n=60 evaluable) |
|---|---|---|
| Sensitivity | 0.55 (0.38-0.71) | 0.57 (0.36-0.76) |
| Specificity | 1.00 (0.90-1.00) | 1.00 (0.91-1.00) |
| Positive predictive value | 0.88 (0.71-0.96) | 0.86 (0.65-0.96) |
| Negative predictive value | 0.97 (0.87-1.00) | 0.98 (0.88-1.00) |
| **Adjudication of the 73 sampled charts** | | |
| Definite dental evaluation or pathology | 24 | |
| Probable | 8 | |
| Absent | 37 | |
| Indeterminate | 4 | |

Values are design-corrected estimates (95% CIs) computed with inverse sampling weights. Indeterminate charts were excluded from the corresponding evaluable denominator, which is why the 2 rules have different denominators. Sampling fractions ranged from 1.0 in the procedure-flagged stratum to 0.0007 in the unflagged stratum, giving an effective sample size of approximately 17 for population-weighted estimates, stratum-specific inference about unflagged patients rests on 12 charts. Adjudication was performed by a single reviewer, blinded to all downstream outcomes by construction.

**eTable 6. Data-Quality and Measurement Checks**

| Item | Finding and implication |
|---|---|
| Microbiologic ascertainment | The SAB and comparator cohorts were identified from prespecified organism-specific ICD-10-CM diagnosis codes, consistent with standard practice in large EHR database studies. These codes constituted the study definitions. |
| Species-level microbiology | Species-level confirmation was unavailable. The A40.8 cohort is therefore reported as code-defined other streptococcal sepsis, not as a species-defined viridans-group cohort. |
| Procedure-table completeness | Operative dental surgery was incompletely represented in structured procedures: 19 454 operative notes describing full-mouth extraction had no matching structured procedure event. The text arm of the definition is therefore mandatory, and procedure-only phenotypes would undercount severely. |
| Imaging dates | Level-1 study dates were 99.6% null. Dates recovered from raw epoch values agreed with independently linked series dates exactly in 99.58% of 5 063 938 linked report-series pairs, within 1 day in 99.93%, and within 7 days in 99.96%. Recovered dates were judged adequate for windowing. |
| Code collisions | Obstetric extraction, laryngeal procedures, skeletal-survey imaging, odontoid imaging, and an alpha-fetoprotein laboratory collision were identified and removed before analysis, superseded counts (61 extraction-flagged and 145 any-signal patients) were replaced before analysis; the pre-removal values are documented in eMethods 1.6 and do not contribute to the reported results. |
| Population denominator | Outcome and Table 1 analyses use the adult 2% sample of 81 945 patients (96 433 before age filtering), with bacteremia codes of any qualifying organism at any time excluded at sampling. |

*eTable 6 (continued)*

| Item | Finding and implication |
|---|---|
| Population comorbidity window | The population pseudo-index is the first qualifying encounter (encounter-based proxy for first health care contact), so no pre-contact history exists by construction and an identical 365-day look-back returned zero events for every covariate. The population comorbidity window therefore spans days −365 to +30, those contrasts are contextual only and were never used for adjustment. |
| Unit of analysis | Among 19 969 patients with an SAB trigger there were 62 567 diagnosis rows and 39 623 distinct patient-dates, and 76.1% of within-patient inter-date gaps were 13 days or less, indicating repeat coding of the same episode. Unique patients, not diagnosis rows, are the unit of analysis throughout. |
| Chart review | Adjudication used a single reviewer, a second independent reader was not available. Interrater reliability could not be estimated. |
| Timing-date capture | First-finding dates were ascertainable for 820 of 1045 patients with documented disease (78.5%): 437 of 528 SAB, 108 of 179 *E coli*, 211 of 250 other streptococcal, and 64 of 88 enterococcal. The remainder had first-ever signals predating the window. Capture is differential across organisms, so cross-organism timing contrasts carry that caveat. |
| Population outcome ascertainment | The locked-definition numerator uses a separate outcome count built in the ascertainment window anchored on the fixed population index (408/81 945, 0.50%), the pathology-only numerator uses a separate outcome count built the same way and joined to the same fixed index (399/81 945, 0.49%). |
| Phenotype implementation | The frozen definition was implemented with exact code matching: ICD-9 extraction codes 23.09/23.19/23.11 had zero yield in the typed procedure table and were dropped (no numerical effect), procedure-description text fallback could not be implemented in the coded-procedure data table and was omitted (strictly narrows the procedure arm), CDT codes were included only when the code produced matching records. |
| Same-day ties | Patients with more than 1 organism on the same earliest date (494 adults, 1.0%) were assigned by code order, excluding them left the unadjusted OR at 4.65 (3.91-5.51). |
| Note-count proxy | Documentation bands count signed/addendum notes, component and Table 3 denominators use 2 or more such notes rather than the earlier rule counting patients with at least one note. |
| Diagnosis-code cohort definition | Cohort eligibility was determined by prespecified organism-specific ICD-10-CM diagnosis codes, consistent with standard practice in large EHR database studies. As in any diagnosis-code–based study, some diagnostic misclassification is possible. |
| Interpretation of a negative result | A negative phenotype indicates that dental disease was not documented, not that dental disease was absent. |

#### eTable 7. Prevalence of Individual Dental Findings and Any Pathology-Only Dental Disease in SAB by Baseline Characteristic

| Characteristic | No. | Abscess No. (%) | Periapical No. (%) | Periodontal No. (%) | Caries No. (%) | Roots No. (%) | Odontogenic No. (%) | Other No. (%) | Any No. (%) |
|---|---|---|---|---|---|---|---|---|---|
| **All patients** | | | | | | | | | |
| All SAB | 16,435 | 221 (1.3) | 135 (0.8) | 137 (0.8) | 122 (0.7) | 66 (0.4) | 113 (0.7) | 59 (0.4) | 517 (3.1) |
| **Age group** | | | | | | | | | |
| Age 18-49 | 5,164 | 130 (2.5) | 67 (1.3) | 59 (1.1) | 86 (1.7) | 43 (0.8) | 50 (1.0) | 45 (0.9) | 273 (5.3) |
| Age 50-64 | 4,644 | 54 (1.2) | 46 (1.0) | 42 (0.9) | 28 (0.6) | 13 (0.3) | 35 (0.8) | 11 (0.2) | 139 (3.0) |
| Age 65-74 | 3,318 | 18 (0.5) | 13 (0.4) | 20 (0.6) | — | — | 14 (0.4) | — | 60 (1.8) |
| Age 75+ | 3,309 | 19 (0.6) | — | 16 (0.5) | — | — | 14 (0.4) | — | 45 (1.4) |
| **Sex** | | | | | | | | | |
| Male | 10,213 | 122 (1.2) | 68 (0.7) | 79 (0.8) | 61 (0.6) | 32 (0.3) | 71 (0.7) | 30 (0.3) | 284 (2.8) |
| Female | 6,195 | 98 (1.6) | 66 (1.1) | 56 (0.9) | 60 (1.0) | 33 (0.5) | 42 (0.7) | 28 (0.5) | 230 (3.7) |
| **Race** | | | | | | | | | |
| White | 12,095 | 182 (1.5) | 99 (0.8) | 99 (0.8) | 108 (0.9) | 51 (0.4) | 83 (0.7) | 50 (0.4) | 400 (3.3) |
| Black | 2,281 | 18 (0.8) | 18 (0.8) | 21 (0.9) | — | — | 16 (0.7) | — | 61 (2.7) |
| Asian | 510 | — | — | — | — | — | — | — | 22 (4.3) |
| Other/unknown race | 1,549 | 13 (0.8) | 12 (0.8) | 11 (0.7) | — | — | — | — | 34 (2.2) |
| **Documentation intensity** | | | | | | | | | |
| 0-1 notes | 2,050 | 0 (0.0) | 0 (0.0) | 0 (0.0) | 0 (0.0) | 0 (0.0) | 0 (0.0) | 0 (0.0) | 0 (0.0) |
| 2-9 notes | 451 | — | 0 (0.0) | — | — | — | 0 (0.0) | — | — |
| 10+ notes | 13,934 | 220 (1.6) | 135 (1.0) | 136 (1.0) | 121 (0.9) | 65 (0.5) | 113 (0.8) | 58 (0.4) | 515 (3.7) |
| **Comorbidity (365 days before index)** | | | | | | | | | |
| Diabetes | 4,177 | 31 (0.7) | 19 (0.5) | 22 (0.5) | 12 (0.3) | — | 20 (0.5) | — | 80 (1.9) |
| Chronic kidney disease | 1,937 | 17 (0.9) | 18 (0.9) | 17 (0.9) | — | — | 14 (0.7) | — | 47 (2.4) |
| End-stage kidney disease | 750 | — | — | — | — | — | — | — | 18 (2.4) |
| Liver disease | 978 | — | — | — | — | — | — | — | 24 (2.5) |
| COPD | 1,615 | 15 (0.9) | — | — | — | — | — | — | 31 (1.9) |
| Malignancy | 2,011 | 14 (0.7) | 14 (0.7) | 12 (0.6) | — | — | — | — | 36 (1.8) |
| HIV or immunocompromise | 633 | — | — | — | — | — | — | — | 14 (2.2) |
| Obesity | 1,224 | 13 (1.1) | 11 (0.9) | — | — | — | — | — | 34 (2.8) |
| Heart failure | 2,195 | 16 (0.7) | 22 (1.0) | 19 (0.9) | 13 (0.6) | 12 (0.5) | 15 (0.7) | — | 63 (2.9) |
| Coronary artery disease | 2,425 | 23 (0.9) | 15 (0.6) | 17 (0.7) | — | — | 11 (0.5) | — | 51 (2.1) |
| Cerebrovascular disease | 1,171 | — | — | 11 (0.9) | — | — | — | — | 29 (2.5) |
| Peripheral vascular disease | 1,599 | 13 (0.8) | — | — | — | — | — | — | 31 (1.9) |
| Native valve disease | 1,111 | 18 (1.6) | 18 (1.6) | 16 (1.4) | 22 (2.0) | 13 (1.2) | 12 (1.1) | — | 53 (4.8) |
| Prosthetic valve | 319 | — | — | — | — | — | — | — | 16 (5.0) |
| Cardiac device | 589 | — | — | — | — | — | — | — | 18 (3.1) |
| Prior infective endocarditis | 379 | 22 (5.8) | 11 (2.9) | — | 20 (5.3) | — | — | — | 37 (9.8) |
| Dementia | 518 | — | — | — | 0 (0.0) | — | — | 0 (0.0) | — |
| Opioid use disorder | 853 | 27 (3.2) | 16 (1.9) | 13 (1.5) | 30 (3.5) | 14 (1.6) | — | — | 53 (6.2) |
| Injection drug use proxy | 1,045 | 27 (2.6) | 17 (1.6) | 13 (1.2) | 31 (3.0) | 14 (1.3) | — | — | 55 (5.3) |
| Vascular catheter codes | 1,296 | 17 (1.3) | 18 (1.4) | 12 (0.9) | 12 (0.9) | 12 (0.9) | — | — | 41 (3.2) |

Values are No. (%) of patients with SAB with the row characteristic who had the column finding documented in days −7 to +30. The Any column uses the pathology-only definition and excludes procedure signals. — indicates a cell with 1 to 10 events (suppressed). Cells with zero events are shown as 0. Findings overlap within patients and do not sum to Any dental disease. Sex was not recorded for 27 patients, so the sex rows sum to 16 408 rather than 16 435. Comorbidities were ascertained from diagnosis codes during the 365 days before the index date on the SAB index, members with a different earliest-organism index were excluded to match **Table 1**. Documentation bands count signed/addendum notes. Abbreviations: SAB, *Staphylococcus aureus* bacteremia.

#### eTable 8. Stratified and Organism-Subtype Sensitivity Analyses

| Analysis | Stratum | SAB No./total (%) | *E coli* BSI No./total (%) | OR (95% CI) |
|---|---|---|---|---|
| **A. Injection drug use proxy** | | | | |
| | Present | 57/1045 (5.45) | 12/448 (2.68) | 2.10 (1.11-3.95) |
| | Absent | 471/15 390 (3.06) | 167/24 494 (0.68) | 4.60 (3.85-5.49) |
| | Pooled (Mantel-Haenszel) | — | — | 4.32 |
| **B. Opioid use disorder** | | | | |
| | Present | 55/853 (6.45) | 7/286 (2.45) | 2.75 (1.24-6.10) |
| | Absent | 473/15 582 (3.04) | 172/24 656 (0.70) | 4.46 (3.74-5.31) |
| | Pooled (Mantel-Haenszel) | — | — | 4.34 |
| **C. Methicillin susceptibility of the index SAB code** | | | | |
| | MSSA only (A41.01) | 279/8930 (3.12) | — | 0.96 (0.80-1.14)[a] |
| | MRSA only (A41.02) | 241/7380 (3.27) | — | [reference] |
| | Both codes at index | 8/125 (6.40) | — | — |

Abbreviations: BSI, bloodstream infection, CI, confidence interval, MRSA, methicillin-resistant *Staphylococcus aureus*, MSSA, methicillin-susceptible *S aureus*, OR, odds ratio, SAB, *S aureus* bacteremia. All analyses use the locked dental definition and the adult earliest-organism cohorts. Panels A and B stratify the primary comparison by a prespecified comorbidity ascertained during the 365 days before index, the association persists in both strata, and the stratum without the comorbidity reproduces the overall unadjusted estimate (4.59, 95% CI, 3.87-5.45), indicating that the excess is not generated by these populations. Mantel-Haenszel estimates pool the 2 strata. Panel C partitions the SAB cohort by the methicillin-susceptibility code recorded at the index date, every adult in the SAB cohort carried A41.01, A41.02, or both, and 125 carried both. [a]MSSA-only versus MRSA-only, indicating no material difference in documented dental disease by methicillin susceptibility.

### Supplementary Figures

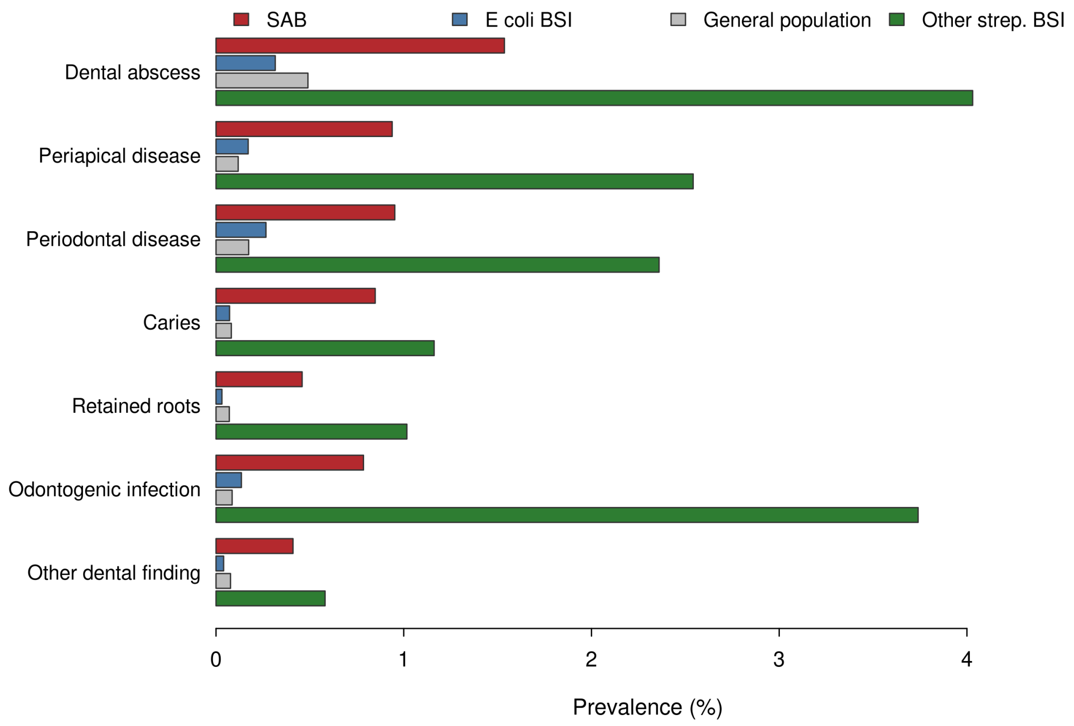


**eFigure 1. Prevalence of individual dental findings by cohort.** Bars show the prevalence of each prespecified dental finding in SAB, *E coli* bloodstream infection (BSI), the general population, and other streptococcal BSI. Denominators are patients with 2 or more signed/addendum notes in the ascertainment window (SAB, n=14 385, *E coli* BSI, n=22 174, general population, n=46 556, other streptococcal BSI, n=2754). Findings overlap within patients and do not sum to the composite outcome. SAB indicates *Staphylococcus aureus* bacteremia.

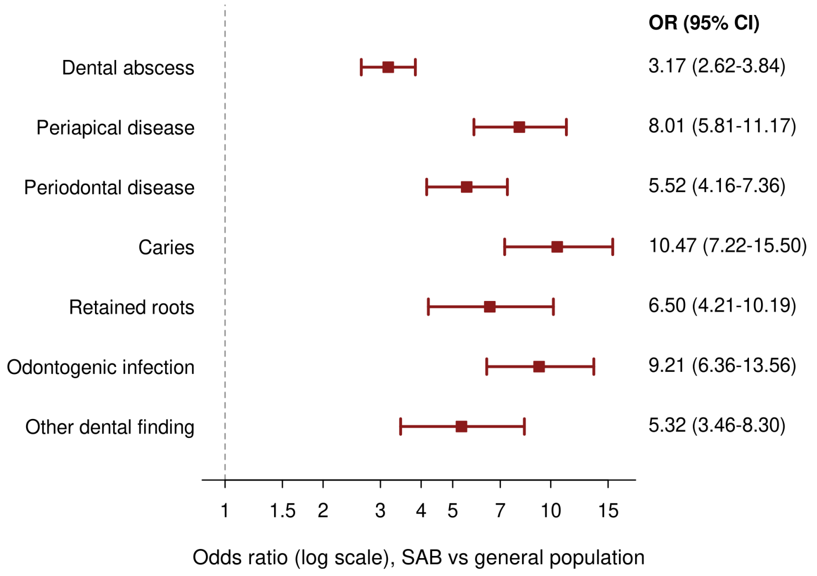


**eFigure 2. Finding-specific odds ratios, SAB vs the general population.** Odds ratios (ORs) are shown with 95% CIs on a logarithmic scale, the dashed line marks the null. Denominators are patients with 2 or more signed/addendum notes in the ascertainment window. All comparisons had $P < .001$ (2-sided Fisher exact test). SAB indicates *Staphylococcus aureus* bacteremia.

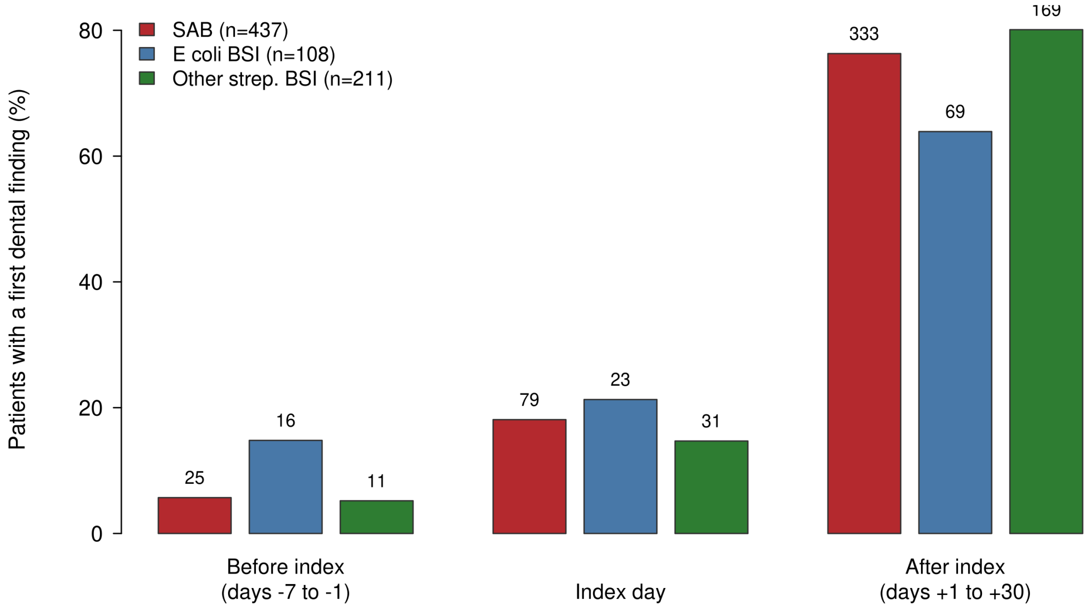


**eFigure 3. Timing of the first dental finding relative to bacteremia, by cohort.** Bars show the percentage of patients with a documented dental finding whose first finding was recorded before the index date, on the index date, or after the index date, counts appear above each bar. Denominators are patients with an ascertainable first-finding date (SAB, n=437, *E coli* bloodstream infection [BSI], n=108, other streptococcal BSI, n=211). SAB indicates *Staphylococcus aureus* bacteremia.

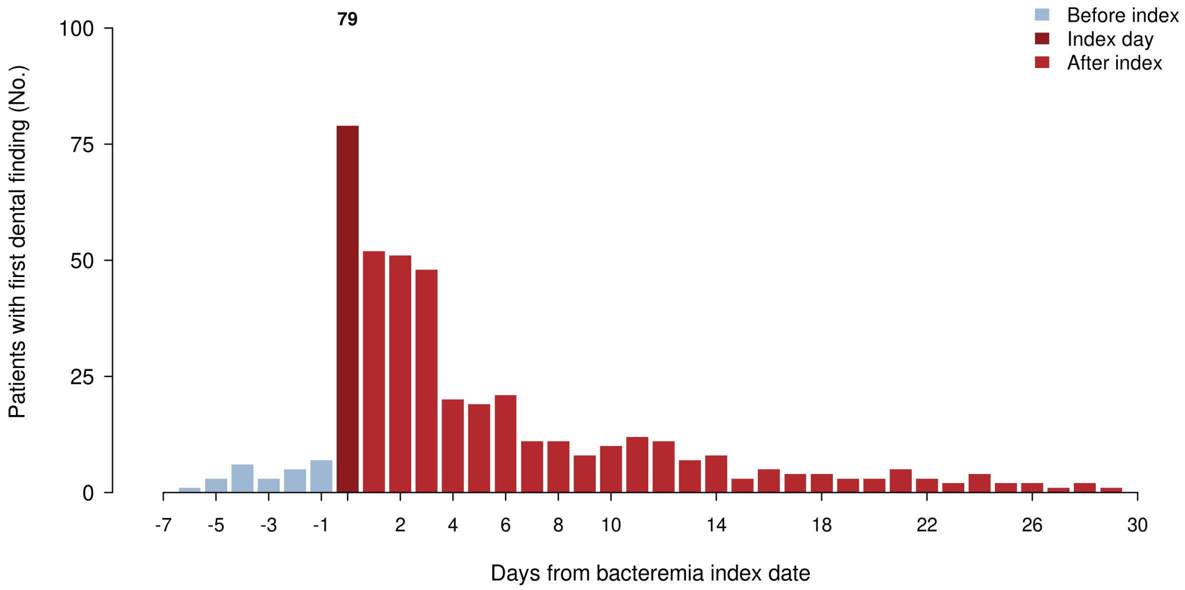


**eFigure 4. Daily distribution of first dental findings in SAB.** Bars show the number of patients with SAB whose first documented dental finding fell on each day from 7 days before through 30 days after the index date (n=437). The distribution peaks on the index date (n=79) and decays thereafter, the median is 3 days after index (interquartile range, 1-8 days). First documentation is not disease onset. SAB indicates *Staphylococcus aureus* bacteremia.

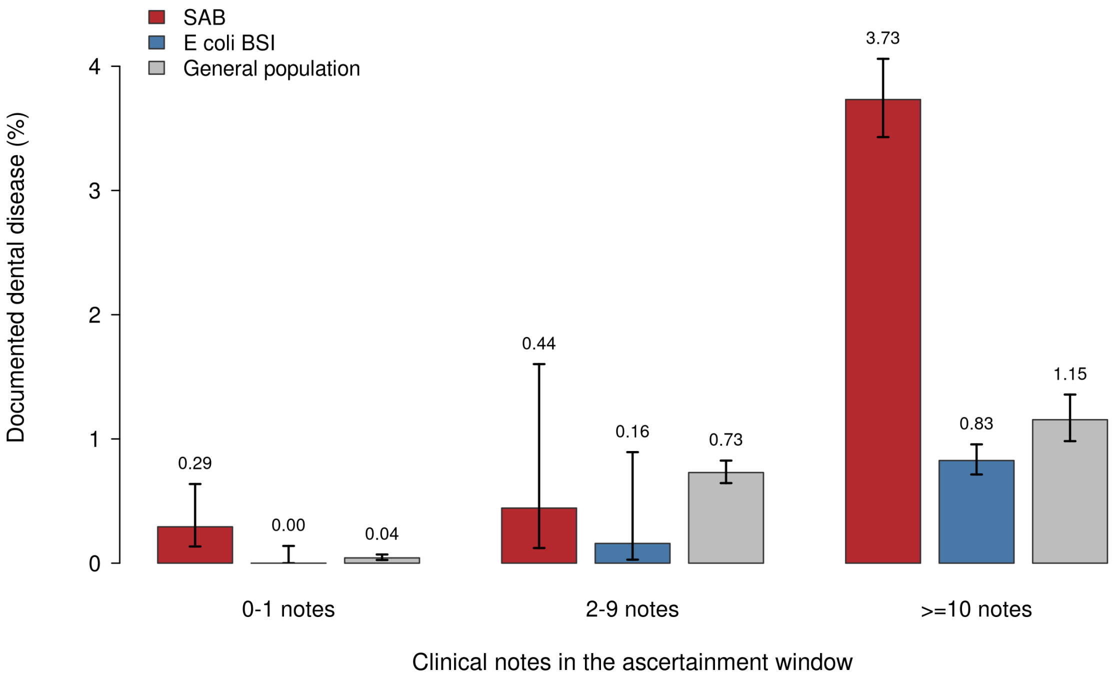


**eFigure 5. Documented dental disease by documentation intensity.** Patients were grouped by the number of cleaned clinical notes in the ascertainment window. Bars show prevalence with 95% Wilson confidence intervals, values above each bar are percentages. Denominators are SAB, 2050/451/13 934, *E coli* bloodstream infection (BSI), 2768/630/21 544, and general population, 35 389/33 999/12 557 for the 0-1, 2-9, and $\geq$10 note bands, respectively. General-population band numerators are 15/248/145 (locked definition). SAB indicates *Staphylococcus aureus* bacteremia.

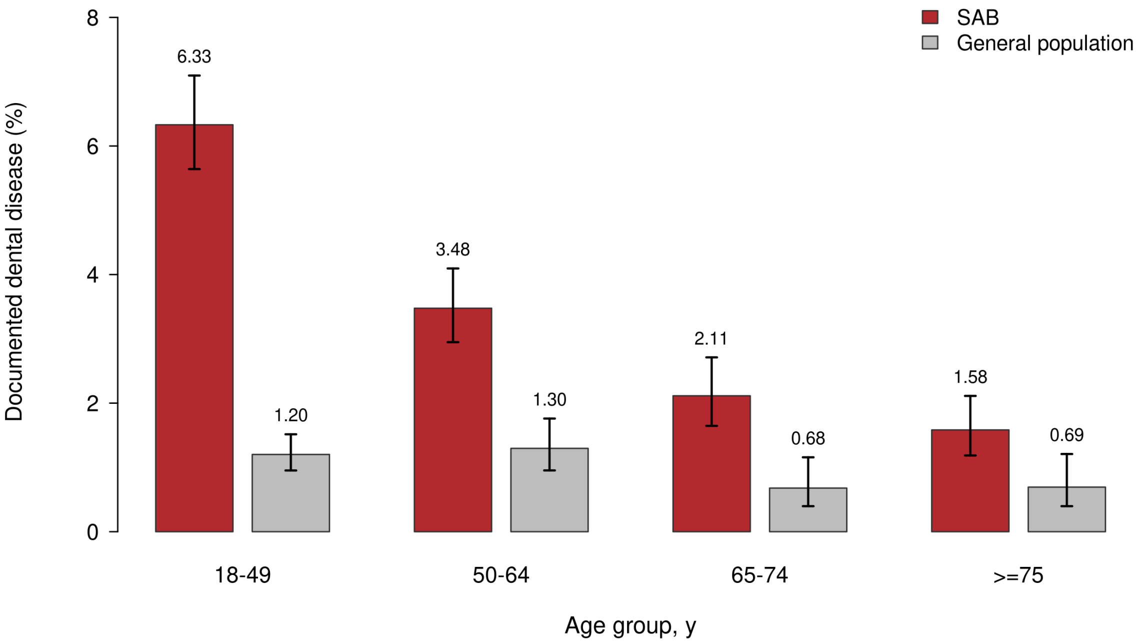


**eFigure 6. Age-stratified prevalence, SAB vs the general population.** Bars show pathology-only prevalence with 95% Wilson confidence intervals within the 10-or-more-note documentation band, in which ascertainment opportunity is most nearly comparable. Values above each bar are percentages. Denominators are SAB, 4313/3941/2838/2842, and general population, 5825/3086/1916/1730, for ages 18-49, 50-64, 65-74, and 75 or older, respectively. The SAB excess narrows with age but persists in every stratum. SAB indicates *Staphylococcus aureus* bacteremia.

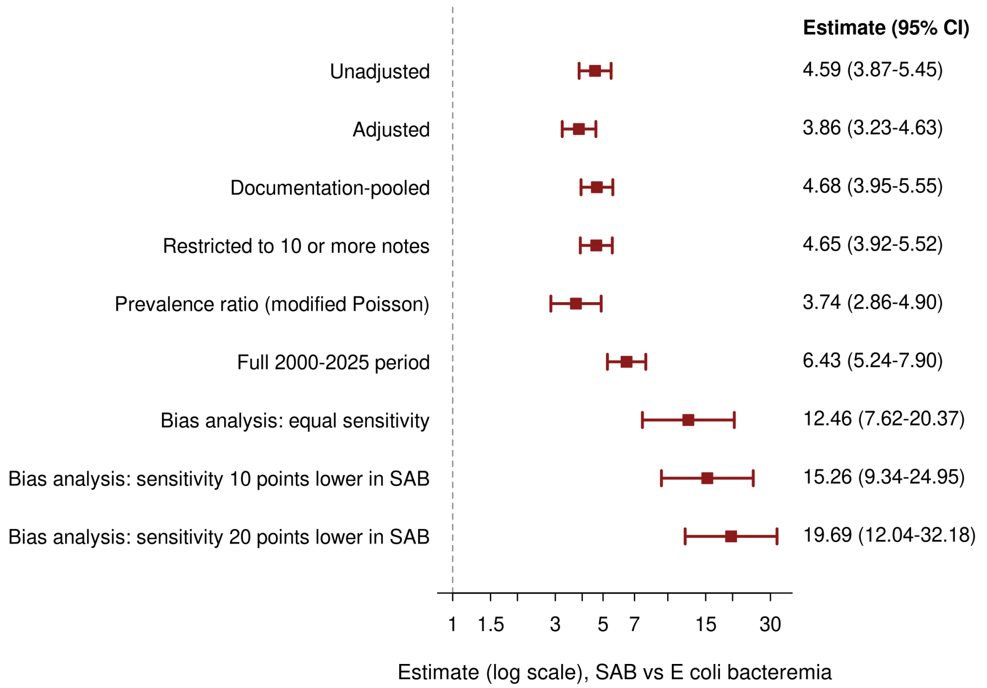


**eFigure 7. Prespecified sensitivity and bias analyses.** Estimates are shown with 95% CIs on a logarithmic scale, the dashed line marks the null. The first 6 rows are odds ratios or the prevalence ratio for SAB vs *E coli* bacteremia (including the full-period sensitivity), the final 3 rows are Rogan-Gladen bias-corrected prevalence ratios with specificity fixed at 0.995 and sensitivity set equal across cohorts or 10 or 20 percentage points lower in SAB. Correction inflates the ratio because the comparator prevalence lies near the false-positive rate. SAB indicates *Staphylococcus aureus* bacteremia.

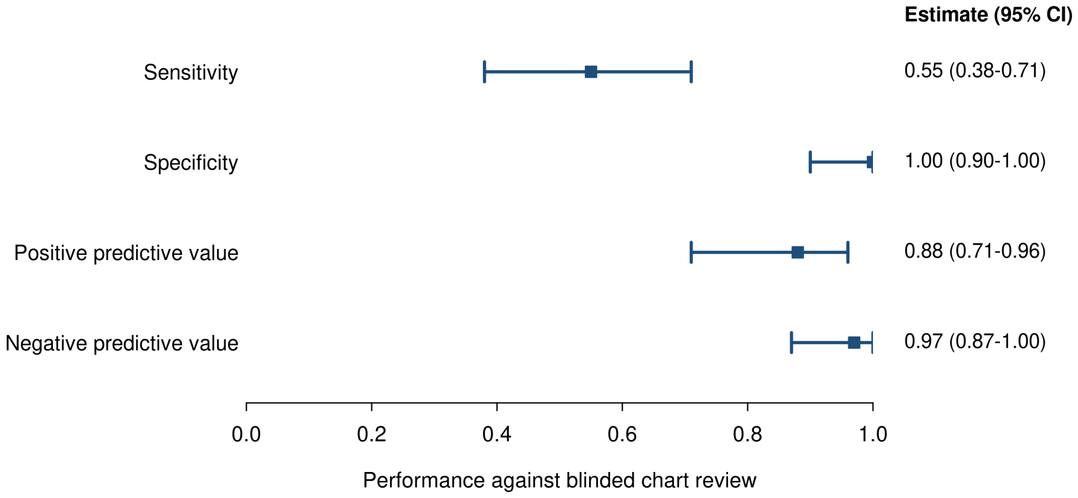


**eFigure 8. Performance of the dental definition against blinded chart review.** Design-corrected sensitivity, specificity, positive predictive value, and negative predictive value are shown with 95% CIs for the dental evaluation rule (n=69 evaluable charts of 73 sampled). Estimates were computed with inverse sampling weights across 5 signal strata plus 1 unflagged stratum (6 total categories).
